# Blood methylation biomarkers are associated with diabetic kidney disease progression in type 1 diabetes

**DOI:** 10.1101/2024.11.28.24318055

**Authors:** Anna Syreeni, Emma H. Dahlström, Laura J. Smyth, Claire Hill, Stefan Mutter, Yogesh Gupta, Valma Harjutsalo, Zhuo Chen, Rama Natarajan, Andrzej S. Krolewski, Joel N. Hirschhorn, Jose C. Florez, GENIE consortium, Alexander P. Maxwell, Per-Henrik Groop, Amy Jayne McKnight, Niina Sandholm, the FinnDiane Study Group

**Affiliations:** Folkhälsan Research Center, Helsinki, Finland; Department of Nephrology, University of Helsinki and Helsinki University Hospital, Helsinki, Finland; Research Program for Clinical and Molecular Metabolism, Faculty of Medicine, University of Helsinki, Helsinki, Finland; Molecular Epidemiology Research Group, Centre for Public Health, Queen’s University Belfast, Belfast, UK; Department of Diabetes Complications and Metabolism, Arthur Riggs Diabetes & Metabolism Research Institute and Beckman Research Institute of City of Hope; Duarte, CA, 91010, USA; Section on Genetics and Epidemiology, Research Division, Joslin Diabetes Center; Boston, MA, 02215, USA; Department of Medicine, Harvard Medical School; Boston, MA, 02215, USA; Programs in Metabolism and Medical & Population Genetics, Broad Institute, Cambridge, MA, USA; Division of Endocrinology and Center for Basic and Translational Obesity Research, Boston Children’s Hospital, Boston, MA, USA; Department of Pediatrics and Genetics, Harvard Medical School, Boston, MA, USA; Diabetes Unit and Center for Genomic Medicine, Massachusetts General Hospital, Boston, MA, USA; Department of Diabetes, Central Clinical School, Monash University, Melbourne, Victoria, Australia; Baker Heart and Diabetes Institute, Melbourne, VIC, Australia

**Author notes:** Corresponding authors:* **DSc Niina Sandholm,**, **Prof Amy Jayne McKnight**.

## Abstract

**Background:** DNA methylation differences are associated with kidney function and diabetic kidney disease (DKD), but prospective studies are scarce. Therefore, we aimed to study DNA methylation in a prospective setting in the Finnish Diabetic Nephropathy Study type 1 diabetes (T1D) cohort.

**Methods:** We analysed baseline blood sample-derived DNA methylation (Illumina’s EPIC array) of 403 individuals with normal albumin excretion rate (early progression group) and 373 individuals with severe albuminuria (late progression group) and followed-up their DKD progression defined as decrease in eGFR to <60 mL/min/1.73m^2^ (early DKD progression group; median follow-up 13.1 years) or end-stage kidney disease (ESKD) (late DKD progression group; median follow-up 8.4 years). We conducted two epigenome-wide association studies (EWASs) on DKD progression and sought methylation quantitative trait loci (meQTLs) for the lead CpGs to estimate genetic contribution.

**Results:** Altogether, 14 methylation sites were associated with DKD progression (*P*<9.4×10^−8^). Methylation at cg01730944 near *CDKN1C* and at other CpGs associated with early DKD progression were not correlated with baseline eGFR, whereas late progression CpGs were strongly associated. Importantly, 13 of 14 CpGs could be linked to a gene showing differential expression in DKD or chronic kidney disease. Higher methylation at the lead CpG cg17944885, a frequent finding in eGFR EWASs, was associated with ESKD risk (HR [95% CI] = 2.15 [1.79, 2.58]). Additionally, we replicated meQTLs for cg17944885 and identified ten novel meQTL variants for other CpGs. Furthermore, survival models including the significant CpG sites showed increased predictive performance on top of clinical risk factors.

**Conclusions:** Our EWAS on early DKD progression identified a podocyte-specific *CDKN1C* locus. EWAS on late progression proposed novel CpGs for ESKD risk and confirmed previously known sites for kidney function. Since DNA methylation signals could improve disease course prediction, a combination of blood-derived methylation sites could serve as a potential prognostic biomarker.

## BACKGROUND

Diabetic kidney disease (DKD) is a devastating complication of diabetes. One-third of individuals with type 1 diabetes (T1D) and severe albuminuria develop end-stage kidney disease (ESKD).^1^ Genetic variability affects the risk of DKD^2,3^ but recent studies highlight the role of epigenetics as well.^4^ One common type of epigenetic modification is DNA methylation, *i.e.*, the attachment of a methyl group at cytosine-guanine dinucleotide (CpG), which contributes to the regulation of gene expression. Epigenome-wide association studies (EWASs) with blood-derived methylation data have identified methylation sites associated with DKD^5–8^ and ESKD^9^ in T1D. Additionally, kidney function, assessed by estimated glomerular filtration rate (eGFR), is associated with DNA methylation, both in individuals with^10–12^ and without^13–15^ diabetes. Remarkably, some top findings, such as methylation site cg17944885 located in a zinc finger gene cluster, have replicated across studies in diabetes cohorts, the general population, and, importantly, multiple ethnic groups. Thus, DNA methylation studies may provide both insights into causal disease pathways and robust prognostic biomarkers to identify individuals at risk.

Epigenetic changes may be dynamic, and changes in DNA methylation can represent either the cause or consequence of DKD. Hyperglycemia can alter DNA methylation, and thereby contribute to metabolic memory — the prolonged effect of hyperglycemia on microvascular complications, even years after the improvement of hyperglycemia.^16,17^

Additionally, genetic variation can regulate DNA methylation.^18,19^ Importantly, methylation quantitative trait loci (meQTLs) can be used to infer causality: We recently identified a methylation site in *REV1* as causally linked to DKD in T1D.^7^

In diabetes, a cross-sectional study of 119 individuals showed differential blood DNA methylation at the early and late stage of DKD.^20^ Furthermore, we have previously shown that 21 of 32 DKD-associated CpGs associated with progression to ESKD^7^, and recently, an EWAS on DKD progression to ESKD identified 17 associated CpGs.^21^ However, no EWAS has yet explored CpGs associated with early progression of DKD in T1D. Here, we employed a prospective study setting and analysed baseline DNA methylation as a predictive biomarker of DKD progression both at the early and late stages of DKD in T1D. Additionally, we searched for meQTLs and serum protein associations for our key methylation findings.

## METHODS

### Cohorts

The study participants were from the ongoing multicentre Finnish Diabetic Nephropathy (FinnDiane) Study that is approved by the Ethics Committee of Helsinki University Central Hospital (491/E5/2006, 238/13/03/00/2015, and HUS-3313-2018) and follows the Declaration of Helsinki. At the study visit, after signing an informed consent, the participants complete questionnaires with the attending nurse or physician, and basic anthropometric measurements are taken.^22^ Blood samples are drawn for DNA extraction and, *e.g*., for serum creatinine measurement. Albuminuria classification is based on two of three consecutive 24-hour or timed overnight urine collections.

#### DKD progression

The early DKD progression sub-cohort comprised 403 individuals (**Figure 1)** with T1D duration ≥10 years, normal albumin excretion rate (AER<30 mg/24h or <20 μg/min), and eGFR≥60 mL/min/1.73m^2^. We collected serum creatinine data from baseline visits and medical records until March 10, 2022, converted Jaffe-method measurements to IDMS units (Creatinine_IDMS_=0.953×Creatinine_Jaffe_–7.261), and calculated eGFR using the revised Chronic Kidney Disease - Epidemiology Collaboration formula (CKD-EPI).^23^ Early DKD progression was defined as eGFR<60 mL/min/1.73m^2^. Thus, the follow-up time was years between the baseline visit and the first date of eGFR<60 mL/min/1.73m^2^ or the latest available eGFR data point.

**Figure 1.**
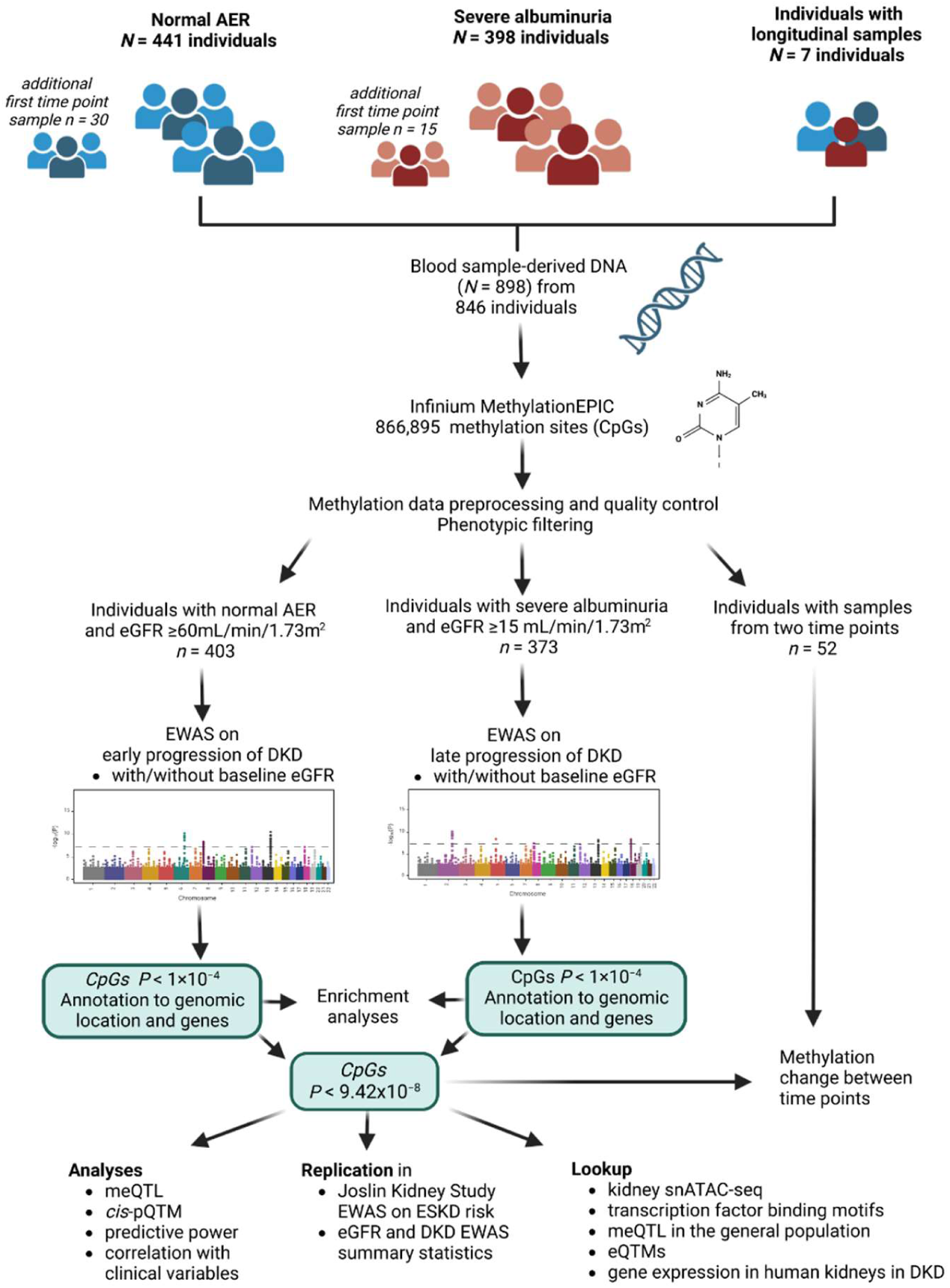
Study setting. Abbreviations: AER=albumin excretion rate; *cis*-pQTM = *cis* protein quantitative trait methylation; DKD=diabetic kidney disease; EWAS=epigenome-wide association study; eGFR=estimated glomerular filtration rate; eQTMs=expression quantitative trait methylations; meQTL=methylation quantitative trait locus; snATAC-seq=single-nucleus transposase-accessible chromatin with sequencing. Created in BioRender. Syreeni, A. (2024) https://BioRender.com/.

The 373 participants in the late DKD progression sub-cohort had T1D and severe albuminuria (AER>300 mg/24h or >200 μg/min) and eGFR>15 mL/min/1.73m^2^, at baseline. We collected data on ESKD, defined as requiring dialysis and/or a transplant, and data on mortality from the Finnish Care Register for Health Care, study visit questionnaires, and medical records. For individuals not yet treated for ESKD, an eGFR record <15 mL/min/1.73m^2^ was considered an ESKD event. The participants were followed up until the event, death, or December 31, 2020.

#### Longitudinal samples

Altogether 52 individuals had DNA samples available at two time points, 3.6–16.4 years apart. Of them, 45 had the second DNA sample analysed as part of the DKD progression cohorts (**Supplemental Figure 1**), whereas seven individuals were new. 30 of 52 individuals had normal AER and eGFR>60 mL/min/1.73m^2^ at both time points. The remaining 22 individuals had normal AER (*n*=8) or moderate albuminuria (*n*=14; AER between 30–300 mg/24h or 20–200 µg/min) at the first time point and progressed to severe albuminuria during follow-up. Additionally, we calculated eGFR slopes between the time points from ≥3 eGFR values ranging over two years.

### DNA methylation assessment

We analysed blood-derived genome-wide DNA methylation with Infinium HD MethylationEPIC v1.0 BeadChip (Illumina, San Diego, CA, USA) within the Northern Ireland Regional Genetics Centre in Belfast. Altogether 798 samples were from our previous cross-sectional DKD EWAS^7^, while 100 were new. The quality control (QC) process details are in **Supplemental Methods**. In brief, from 898 samples and 866,895 methylation probes, one sample and 105,357 probes were removed during QC. Thereafter, we extracted methylation M-values of the remaining 761,538 methylation probes from 897 samples using *’RnBeads’*. Additionally, we calculated principal components (PC) from the non-negative control probe intensities and mean methylation (mean M-value) of probes known to have invariable methylation levels in blood-based DNA.^24^ These variables were used in the subsequent EWASs to correct for technical deviations.

### Statistical analysis

#### DKD progression

We analysed associations between each methylation site and DKD progression separately for the early and late DKD progression cohorts using a Cox proportional-hazards model adjusted for sex, baseline age, six estimated white blood cell counts (WCCs), PCs 1–3, and intrapersonal mean M from invariable sites. The second model included baseline eGFR as an additional covariate. Significance threshold was *P*<9.4×10^−8^, as recommended for the EPIC array.^25^

#### Longitudinal analyses

Using longitudinal data, we compared methylation change (Δmethylation) over time between DKD progressors and non-progressors using logistic regression and residualised methylation values (**Supplemental Methods**). Additionally, we tested the association between eGFR slope (dependent variable) and Δmethylation using linear regression (**Supplemental Methods**).

#### Replication

We included several look-up replication cohorts: United Kingdom and Republic of Ireland (UK-ROI, *n*=372) T1D cohort with DKD EWAS data^7^, Joslin Kidney Study with prospective kidney failure EWAS data (*n*=277)^21^ as well as eGFR-EWAS summary statistics from the Chronic Renal Insufficiency cohort (CRIC)^10^, the Hong Kong diabetes register^11^, and the general population.^13–15^ To assess whether diabetes contributed to the associations, we compared ESKD-DKD (*n*=108) *vs*. ESKD due to other causes (*n*=71)^9^, DKD (*n*=252, UK-ROI) *vs.* individuals without diabetes nor kidney disease (*n*=340, from the Northern Ireland Cohort for the Longitudinal Study of Ageing (NICOLA), and ESKD-DKD (*n*=108, UK-ROI/Renal Transplant Collection samples) *vs*. the 340 NICOLA participants.

#### Sensitivity analyses

We tested the association with baseline eGFR and carried out both 10-year-risk and competing risk analyses regarding late DKD progression. Additionally, we studied pleiotropy with correlation analysis of the methylation data and baseline characteristics (**Supplemental Methods**).

#### Predictive power

We compared the concordance indices (C-index) of Cox models using clinical risk factors, both with and without CpG methylation values. The chosen clinical variables were associated with (early or late) DKD progression in a univariable (*P*<0.25) and multivariable Cox regression models (*P*<0.10). Additionally, we included age, sex, and methylation assay QC-variables in all models, including the clinical model, to separate the methylation effect from technical variability. We compared models: 1) clinical variables, 2) clinical variables and baseline eGFR, and 3) clinical variables, eGFR, and CpG methylation. Additionally, we created a model incorporating all significant CpGs with clinical variables and eGFR to study the cumulative effect. An increase in the C-index (*P*<0.05) was considered significant.

### Annotation of methylation sites

#### CpG location

We examined the overlap of CpG genomic locations with kidney open chromatin peaks^26−28^ utilising the Susztaklab Kidney Biobank^29^, with TF motifs^30,31^, quantitative trait methylation (eQTMs) datasets^27,34–36,37^, and meQTLs^32,33^. We performed a meQTL analysis to identify local (*cis,* ±1 Mb) and distal (*trans*) genetic effects for the CpGs (**Supplemental Methods**).

#### Gene expression in the kidney

Differential gene expression in human diabetic kidneys, was studied in datasets ^38,39,40,41^ collected into the Nephroseq database v5^42^ (**Supplemental Methods**). Additionally, we studied two human DKD kidney tissue gene expression datasets^43,44^, preprocessed similarly to the previous study.^45^ Kidney single-cell gene expression data^46^ were accessed through the Kidney Interactive Transcriptomics database.^47^

#### Protein expression

Serum proteome data measured with OLINK^®^ Ht assay at SciLifeLab in Uppsala were available for 188 individuals with normal AER (main analysis group) and 127 individuals with severe albuminuria (replication group). We analysed the association between methylation and proteins levels of *cis*-located genes (*i.e*., *cis* protein quantitative trait methylation (*cis*-pQTM), **Supplemental Methods)**.

#### Enrichment analysis

We analysed the enrichment of Gene ontology (GO) terms and Kyoto Encyclopedia of Genes and Genomes (KEGG) pathways with the R package *’missMethyl’* (v.1.22.0) *gometh-*function for early and late DKD progression separately. Additionally, we assessed CpG trait enrichment using EWAS Toolkit.^48^

## RESULTS

### CpGs associated with DKD progression

In the early DKD progression cohort of 403 individuals, 37% were women, and mean age was 42 years (**Table 1**). Over the 13.1-year (interquartile range: 8.4–16.9) follow-up, DKD progressed in 49 individuals. EWAS identified two methylation sites significantly associated (*P*<9.4×10^−8^) with early DKD progression: cg25013571 between *PLPBP* and *ADGRA2* (HR [95%CI] = 3.35 [2.18, 5.13]) and cg05831784 in *HAO1* (0.42 [0.30, 0.57]; **Table 2**, **Figure 2, Supplemental Figure 2**). Cg25013571 (*PLPBP/ADGRA2*) remained significant in EWAS adjusted for baseline eGFR, whereby the cg05831784 (*HAO1*) association was modestly attenuated. Furthermore, in eGFR-adjusted EWAS, cg06334496 in *TMEM70* and cg01730944 close to the transcription start site (TSS) of *CDKN1C*, alias *p57^Kip2^*, were significantly associated with early DKD progression. Cg01730944 was generally hypomethylated (beta-values<0.05) (**Figure 3A, Supplemental Figure 3**), and low methylation values were associated with risk of DKD progression (**Figure 3B)**.

**Figure 2.**
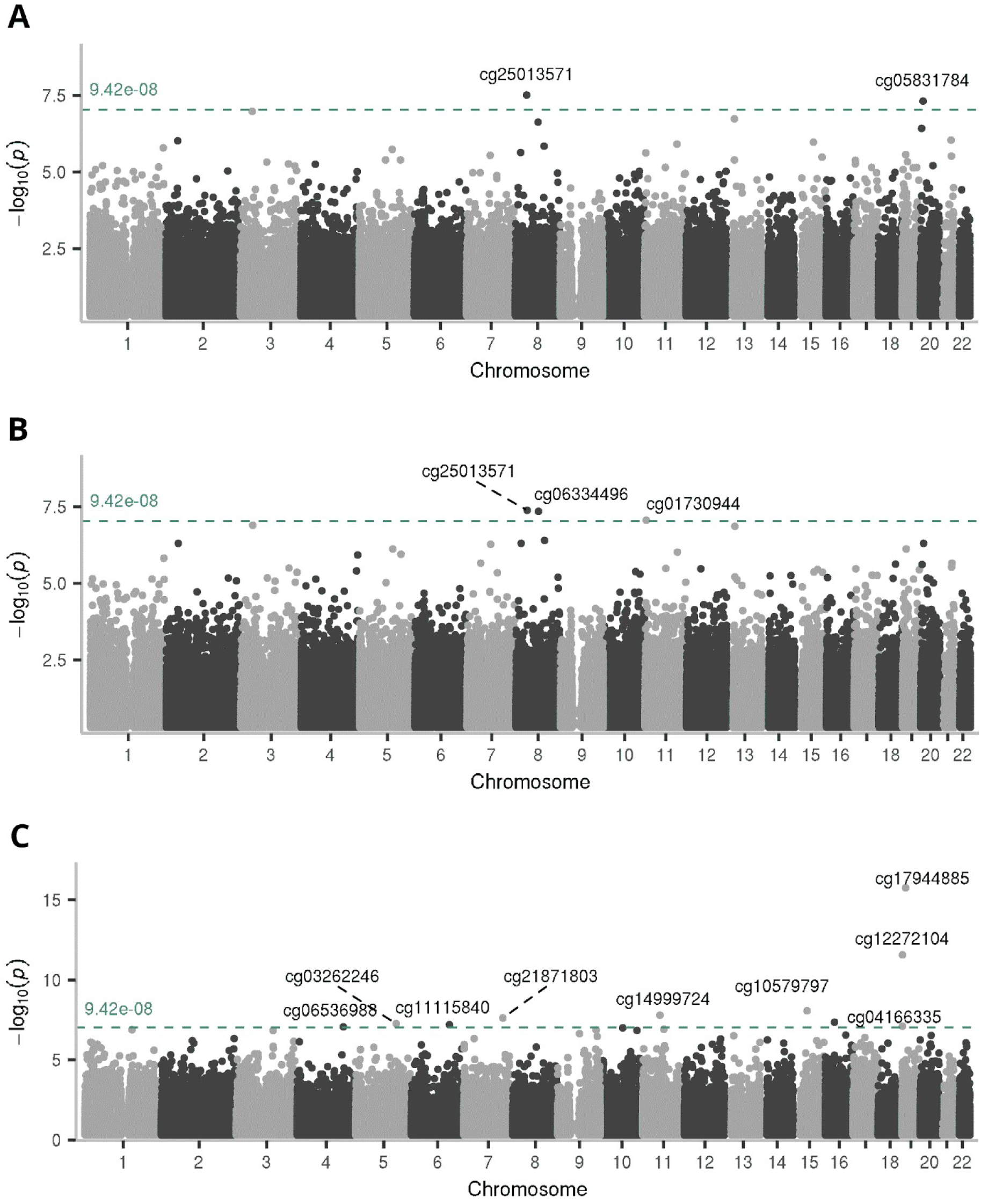
Manhattan plots show the results of EWASs on DKD progression. **A)** Results from the EWAS on early DKD progression, **B)** early DKD progression EWAS additionally adjusted for the baseline eGFR, and **C)** results from the EWAS on late DKD progression (to ESKD). X-axis shows the chromosomal position and y-axis shows the −log_10_ of the association *P*-value. Methylation sites reaching epigenome-wide significance (*P*<9.4×10^−8^, green line) are annotated into the plot.

**Figure 3.**
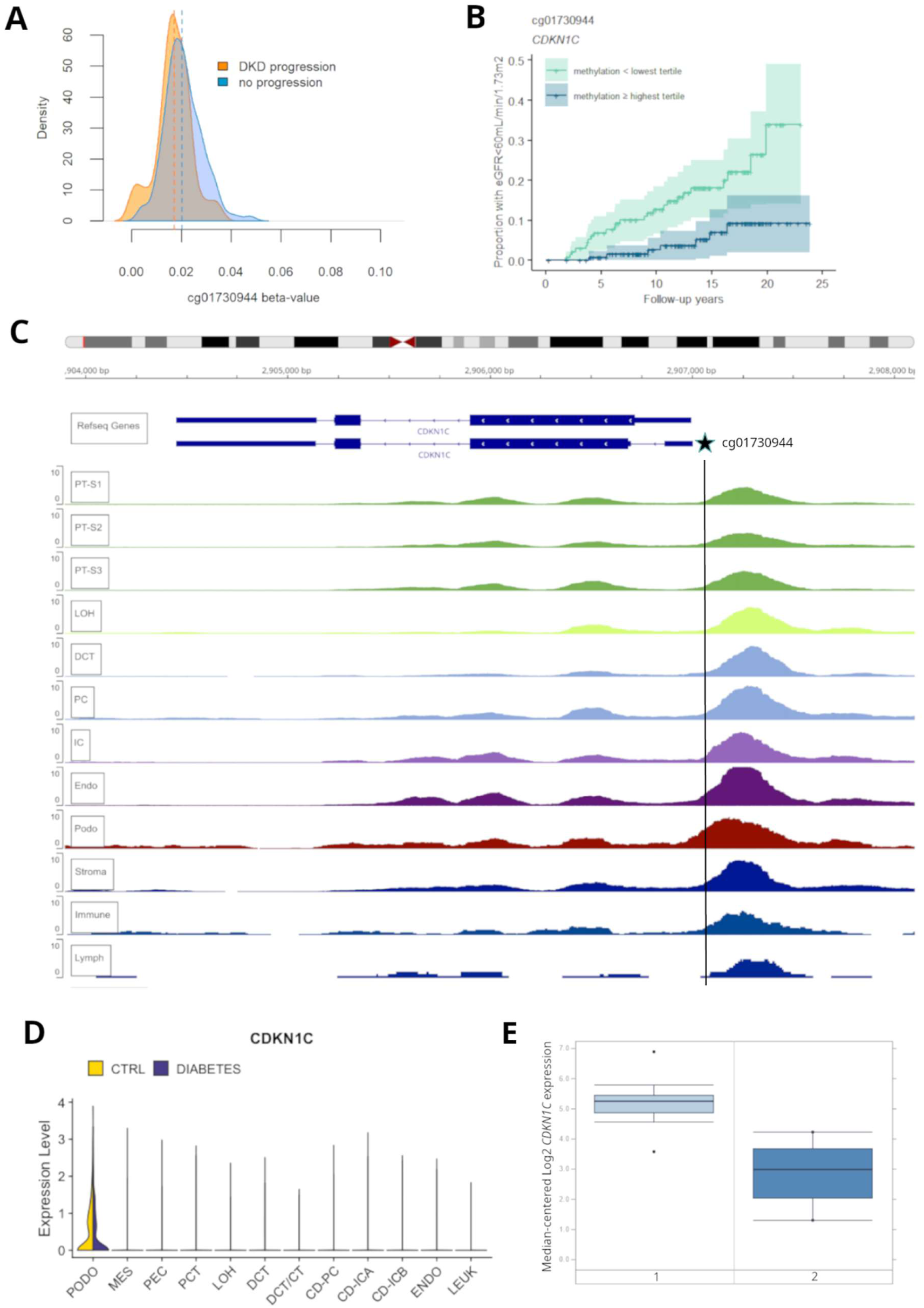
Methylation site cg01730944 is located close to *CDKN1C*. **A)** Density plot of early DKD progression cohort (*n*=403) baseline methylation beta values of cg01730944 shows lower methylation in individuals with progressing DKD during follow-up [eGFR decline <60 mL/min/1.73 m^2^ (in orange)] compared to individuals who do not progress (light blue) **B)** Kaplan–Meier plot compares individuals in the lowest and highest tertile for cg01730944 methylation and shows the proportion of individuals progressing to eGFR<60 mL/min/1.73 m^2^ during follow-up. **C)** Open chromatin peaks in kidney cell types; human kidney single-nucleus transposase-accessible chromatin data (Version 2) on 57,229 cells^27^ accessed in Susztaklab Kidney Biobank.^29^ Figure is adapted from https://susztaklab.com/Human_snATAC/, and cg01730944 position is incorporated. **D)** Kidney single-cell expression data of 23,980 nuclei^46^ shows that *CDKN1C* is mainly expressed in podocytes. Adapted from Humphrey’s Lab browser at http://humphreyslab.com^47^ **E)** *In vivo* expression of *CDKN1C* in human glomerular cells^38^ shows lower expression (fold-change=−4.95, *P*=4.9×10^−5^ in diabetic kidney disease (group 2, *n*=9) compared to individuals without DKD (group 1, *n*=13). Figure adapted from Nephroseq v.5 database^42^ at https://www.nephroseq.org/. Abbreviations: PT-S1–PT-S3=proximal tubule segments 1–3; LOH=loop of Henle; DCT=distal convoluted tubule; PC=principal cells of collecting duct; IC=intercalated cells, Endo=endothelia; Podo=podocytes; Immune=immune cells; lymph=lymphocytes; MES=mesenchyme, PEC=parietal epithelial cell; PCT=proximal convoluted tubule; DCT/CT=distal convoluted tubule/connecting tubule; CD-PC=collecting duct - principal cell; CD-ICA=collecting duct - intercalated cells A; CD-ICB=collecting duct - intercalated cells B; Leuk=leukocytes

**Table 1.** Baseline characteristics of the study participants.

|  | Early DKD progression cohort (n=403) |  | Late DKD progression cohort (n=373) |  |
| --- | --- | --- | --- | --- |
|  | No event | Event (eGFR decline <60) during follow-up | No event | Event (ESKD) during follow-up |
| *n* | 354 | 49 | 167 | 206 |
| Women, *n* (%) | 131 (37) | 20 (41) | 53 (32) | 89 (43) |
| Age, years | 42 ± 11 | 45 ± 14 | 43 ± 12 | 43 ± 10 |
| T1D duration, years | 27 ± 9 | 28 ± 12 | 30 ± 9 | 30 ± 10 |
| Systolic blood pressure, mmHg | 133 ± 18 | 135 ± 18 | 143 ± 17 | 149 ± 21 |
| Diastolic blood pressure, mmHg | 78 ± 8.6 | 78 ± 9.7 | 82 ± 9.9 | 84 ± 11 |
| Pulse pressure, mmHg | 55 ± 15 | 58 ± 19 | 61 ± 16 | 65 ± 19 |
| HbA<sub>1c</sub>, mmol/mol (%) | 66.2 ± 13.6 | 70.2 ± 15.8 | 71.7 ± 16.0 | 75.6 ± 18.0 |
| Central obesity, *n* (%) | 163 (46.4) | 31 (66.0) | 119 (73.0) | 135 (66.5) |
| Triglycerides, mg/dL | 82 (32, 113) | 93 (75, 113) | 113 (82, 165) | 142 (105, 218) |
| Granulocytes, % | 63 (57, 70) | 63 (57, 69) | 67 (60, 73) | 69 (64, 74) |
| Monocytes, % | 7 (5, 9) | 8 (6, 9) | 8 (6, 10) | 8 (6, 9) |
| CD4<sup>+</sup> T-cells, % | 11 (8, 14) | 12 (8, 17) | 10 (7, 13) | 10 (6, 13) |
| CD8<sup>+</sup> T-cells, % | 5 (2, 8) | 3 (1, 8) | 4 (2, 8) | 4 (1, 7) |
| B-cells, % | 4 (2, 5) | 3 (2, 5) | 3 (2, 5) | 2 (0.8, 4) |
| NK-cells, % | 4 (0.1, 7) | 5 (1, 8) | 0.9 (0.0, 4.6) | 0.7 (0.0, 4.3) |
| eGFR, mL/min/1.73 m<sup>2</sup> | 105 ± 14 | 100 ± 19 | 85 (71, 106) | 44 (28, 67) |
| Follow-up time, years | 13.5 (8.9, 17.3) | 9.7 (4.4, 14.4) | 14.1 (7.5, 21.3) | 6.1 (2.9, 10.2) |

Data are expressed as mean ± standard deviation or median (interquartile range)

**Table 2.**
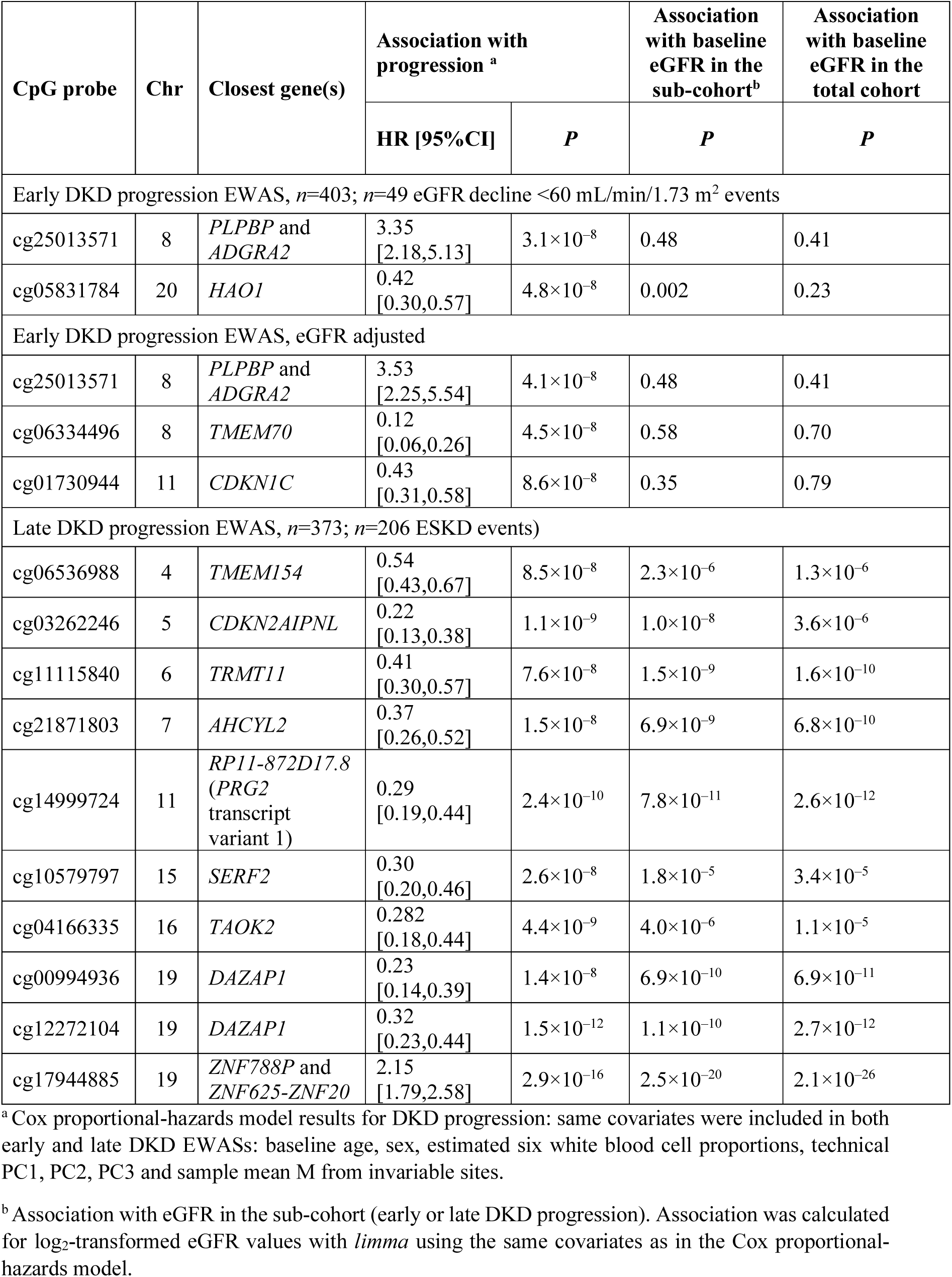
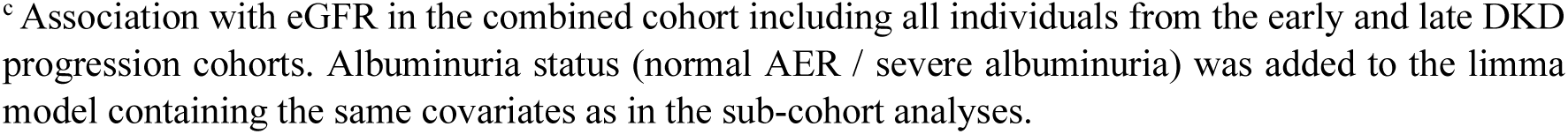
Epigenome-wide significant methylation CpGs sites for the progression kidney disease.

| CpG probe | Chr | Closest gene(s) | Association with progression <sup>a</sup> |  | Association with baseline eGFR in the sub-cohort<sup>b</sup> | Association with baseline eGFR in the total cohort |
| --- | --- | --- | --- | --- | --- | --- |
|  |  |  | HR [95%CI] | *P* | *P* | *P* |
| Early DKD progression EWAS, *n*=403; *n*=49 eGFR decline <60 mL/min/1.73 m<sup>2</sup> events |  |  |  |  |  |  |
| cg25013571 | 8 | *PLPBP* and *ADGRA2* | 3.35 [2.18,5.13] | 3.1×10<sup>-8</sup> | 0.48 | 0.41 |
| cg05831784 | 20 | *HAO1* | 0.42 [0.30,0.57] | 4.8×10<sup>-8</sup> | 0.002 | 0.23 |
| Early DKD progression EWAS, eGFR adjusted |  |  |  |  |  |  |
| cg25013571 | 8 | *PLPBP* and *ADGRA2* | 3.53 [2.25,5.54] | 4.1×10<sup>-8</sup> | 0.48 | 0.41 |
| cg06334496 | 8 | *TMEM70* | 0.12 [0.06,0.26] | 4.5×10<sup>-8</sup> | 0.58 | 0.70 |
| cg01730944 | 11 | *CDKN1C* | 0.43 [0.31,0.58] | 8.6×10<sup>-8</sup> | 0.35 | 0.79 |
| Late DKD progression EWAS, *n*=373; *n*=206 ESKD events) |  |  |  |  |  |  |
| cg06536988 | 4 | *TMEM154* | 0.54 [0.43,0.67] | 8.5×10<sup>-8</sup> | 2.3×10<sup>-6</sup> | 1.3×10<sup>-6</sup> |
| cg03262246 | 5 | *CDKN2AIPNL* | 0.22 [0.13,0.38] | 1.1×10<sup>-9</sup> | 1.0×10<sup>-8</sup> | 3.6×10<sup>-6</sup> |
| cg11115840 | 6 | *TRMT11* | 0.41 [0.30,0.57] | 7.6×10<sup>-8</sup> | 1.5×10<sup>-9</sup> | 1.6×10<sup>-10</sup> |
| cg21871803 | 7 | *AHCYL2* | 0.37 [0.26,0.52] | 1.5×10<sup>-8</sup> | 6.9×10<sup>-9</sup> | 6.8×10<sup>-10</sup> |
| cg14999724 | 11 | *RP11-872D17.8* (PRG2 transcript variant 1) | 0.29 [0.19,0.44] | 2.4×10<sup>-10</sup> | 7.8×10<sup>-11</sup> | 2.6×10<sup>-12</sup> |
| cg10579797 | 15 | *SERF2* | 0.30 [0.20,0.46] | 2.6×10<sup>-8</sup> | 1.8×10<sup>-5</sup> | 3.4×10<sup>-5</sup> |
| cg04166335 | 16 | *TAOK2* | 0.282 [0.18,0.44] | 4.4×10<sup>-9</sup> | 4.0×10<sup>-6</sup> | 1.1×10<sup>-5</sup> |
| cg00994936 | 19 | *DAZAP1* | 0.23 [0.14,0.39] | 1.4×10<sup>-8</sup> | 6.9×10<sup>-10</sup> | 6.9×10<sup>-11</sup> |
| cg12272104 | 19 | *DAZAP1* | 0.32 [0.23,0.44] | 1.5×10<sup>-12</sup> | 1.1×10<sup>-10</sup> | 2.7×10<sup>-12</sup> |
| cg17944885 | 19 | *ZNF788P* and *ZNF625-ZNF20* | 2.15 [1.79,2.58] | 2.9×10<sup>-16</sup> | 2.5×10<sup>-20</sup> | 2.1×10<sup>-26</sup> |

<sup>a</sup> Cox proportional-hazards model results for DKD progression: same covariates were included in both early and late DKD EWASs: baseline age, sex, estimated six white blood cell proportions, technical PC1, PC2, PC3 and sample mean M from invariable sites.
<sup>b</sup> Association with eGFR in the sub-cohort (early or late DKD progression). Association was calculated for log<sub>2</sub>-transformed eGFR values with *limma* using the same covariates as in the Cox proportional-hazards model.
<sup>c</sup> Association with eGFR in the combined cohort including all individuals from the early and late DKD progression cohorts. Albuminuria status (normal AER / severe albuminuria) was added to the limma model containing the same covariates as in the sub-cohort analyses.

The 373 individuals with severe albuminuria at baseline were followed-up for a median of 8.4 (interquartile range: 4.1–15.4) years. Altogether, 38% were women, and mean age 43 years. Individuals (*n*=206, 55%) who developed ESKD had lower baseline eGFR compared to those 167 who did not progress to ESKD (43.5 *vs.* 84.9 mL/min/1.73m^2^, **Table 1**).

EWAS on late DKD progression identified ten significant CpGs (*P*<9.4×10^−8^) from nine genomic loci (**Table 2**). Higher methylation at the top site cg17944885 between *ZNF788P* and *ZNF625*-*ZNF20* (chr19p13.2) was associated with ESKD risk (HR [95%CI] = 2.15 [1.79, 2.58]). The nine additional CpGs exhibited lower methylation as risk for progression of DKD to ESKD (HRs<1.0), supporting the previously suggested trend of general hypomethylation in advanced DKD.^8^ In competing risk analysis (*n*=51 death events), eight CpGs remained significantly associated with ESKD risk (**Supplemental Table 1**).

The top ten CpGs were associated with baseline eGFR (**Table 2**), which likely attenuated their association with ESKD risk in the eGFR-adjusted EWAS (**Supplemental Table 2**), where no epigenome-wide significant associations were seen (**Supplemental Figure 4**).

Longitudinal dataset showed that methylation levels of the 14 DKD progression-associated CpGs seemed relatively stable over time: only at cg17944885 (chr19p13.2) progressors from normal AER to severe albuminuria had increase in methylation, *i.e*., in the expected direction, when compared to non-progressors (*P*=0.049; non-significant after Bonferroni-correction; **Supplemental Figures 5** and **6**). No association between Δmethylation and eGFR slope was observed (**Supplemental Table 3**).

### Replication

We studied several EWAS datasets to validate the lead findings. Notably, the CpGs associated with early DKD progression were not associated with eGFR, implying that EWASs on eGFR are unsuitable for replicating these signals, and no cohort with a comparable early progression phenotype and EWAS data currently exists. Nevertheless, three of four early DKD progression-associated CpGs showed differential methylation in DKD (*n*=252) compared to healthy individuals (*n*=340) without diabetes and kidney disease: cg25013571 (*PLPBP/ADGRA2*), cg05831784 (*HAO1*), and cg01730944 (*CDKN1C*), (*P-*values<1.4×10^−6^, **Supplemental Table 4)**.

Eight of ten late DKD progression-associated CpGs were nominally (*P*<0.05) or significantly (*P<*3.6×10^−3^; Bonferroni correction) associated with eGFR in the replication datasets. Remarkably, higher methylation at cg17944885 (chr19p13.2) was consistently associated with lower eGFR in five eGFR EWASs (*P*<1.4×10^−9^), DKD in the UK-ROI cohort (*P*=9.5×10^−16^), and risk of ESKD in the JKS cohort (*P*<6.2×10^−4^). Additionally, cg00994936 and cg12272104 (*DAZAP1*) were robustly replicated; Cg12272104 is already a known eGFR-associated CpG.^13^ Notably, the novel cg21871803 (*AHCYL2*) was significantly replicated in the eGFR slope EWAS (*P*=1.3×10^−4^)^11^ and nominally in EWAS on DKD progression to ESKD^21^.

### Association with clinical variables

Methylation sites associated with early DKD progression correlated only modestly with baseline clinical variables indicating that methylation at these sites is not strongly affected by these factors (**Supplemental Figure 7**). Nine of ten late DKD progression-associated CpGs correlated with baseline eGFR and only modestly with other clinical variables; for instance, only two sites correlated with HbA_1c_ (**Supplemental Figure 8**). Interestingly, methylation values of late DKD progression-associated CpGs correlated with one another (**Supplemental Figure 9**).

### Prediction of kidney outcomes

When predicting early DKD progression, baseline eGFR did not improve the clinical model: the C-index was 0.783 *vs.* 0.775 (Cox model with clinical variables). This implies that baseline eGFR does not help distinguishing early DKD progressors. The top four CpG sites, separately, did not improve the model (**Supplemental Figure 10)**, whereas a model including all four performed better compared with a model with clinical variables and eGFR (C-index 0.859 *vs.* 0.783, *P*=0.01, **Figure 4**).

**Figure 4.**
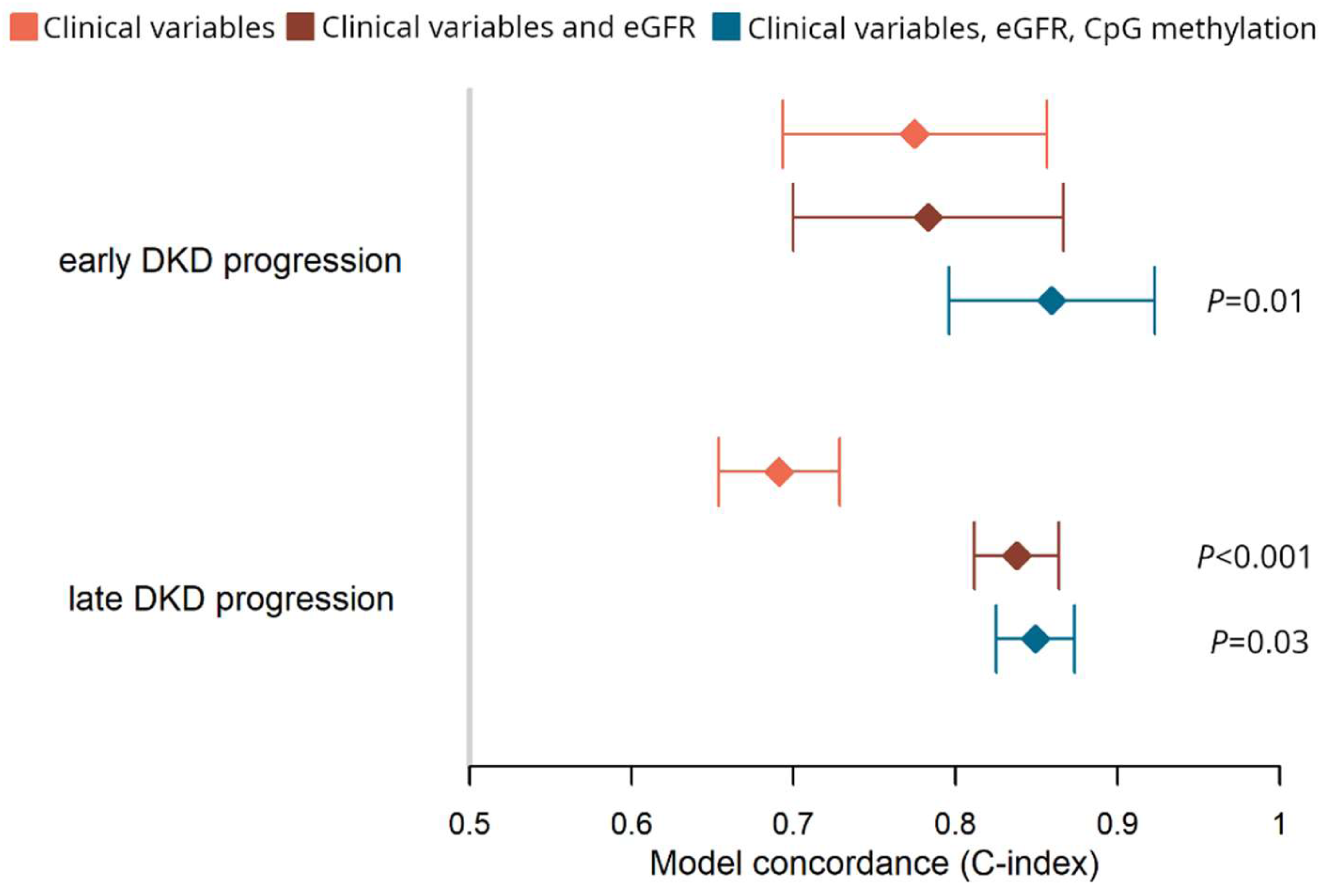
Predictive power of the lead CpGs. The diamonds show the concordance (C-index) and its 95% confidence intervals of three Cox proportional-hazards models applied for the early (*n*=393 with non-missing variables) and late DKD progression (*n*=363 with non-missing variables) cohorts. *P*-values denote the significance of the increase in concordance index compared to the previous model; The significant *P*-values (*P*<0.05) are marked in the figure. The first model, “Clinical variables” (orange color), included baseline triglyceride concentration, central obesity, and current smoking status for the early DKD progression analysis, and triglyceride concentration, HbA_1c_, and systolic blood pressure for the late DKD progression analysis. Additionally, the model included six white blood cell proportions, technical PCs 1–3, mean methylation M value from invariable sites, age, and sex. The second model (red color) included additionally baseline eGFR. The third model included methylation M values for four (early DKD progression-associated: cg25013571, cg05831784, cg06334496, and cg01730944) or nine (late DKD progression-associated: cg06536988, cg03262246, cg11115840, cg21871803, cg14999724, cg10579797, cg04166335, cg12272104, and cg17944885) methylation sites.

As expected, adding baseline eGFR into the clinical model improved the Cox model for late DKD progression (C-index 0.838 *vs.* 0.691, *P*<0.001). The significant CpGs, separately, did not improve the model (**Supplemental Figure 11**) but a model including them all outperformed the clinical model with eGFR (C-index 0.849 *vs.* 0.838, *P*=0.03).

### meQTLs

We subsequently studied the impact of genetic variability on methylation levels at the top sites. We identified nine independent *cis*-meQTLs associated with methylation at seven CpGs (*P*<0.05 at FDR<0.05, **Table 3, Supplemental Table 5**). These included rs555097 for cg14999724 (*RP11-872D17.8*) without prior *cis*-meQTLs in the Genetics of DNA Methylation Consortium (GoDMC) data (**Supplemental Figure 12**). Our lead *cis*-meQTL rs4804653 for cg17944885 (chr19p13.2) was found also in the general population (GoDMC).

**Table 3.**
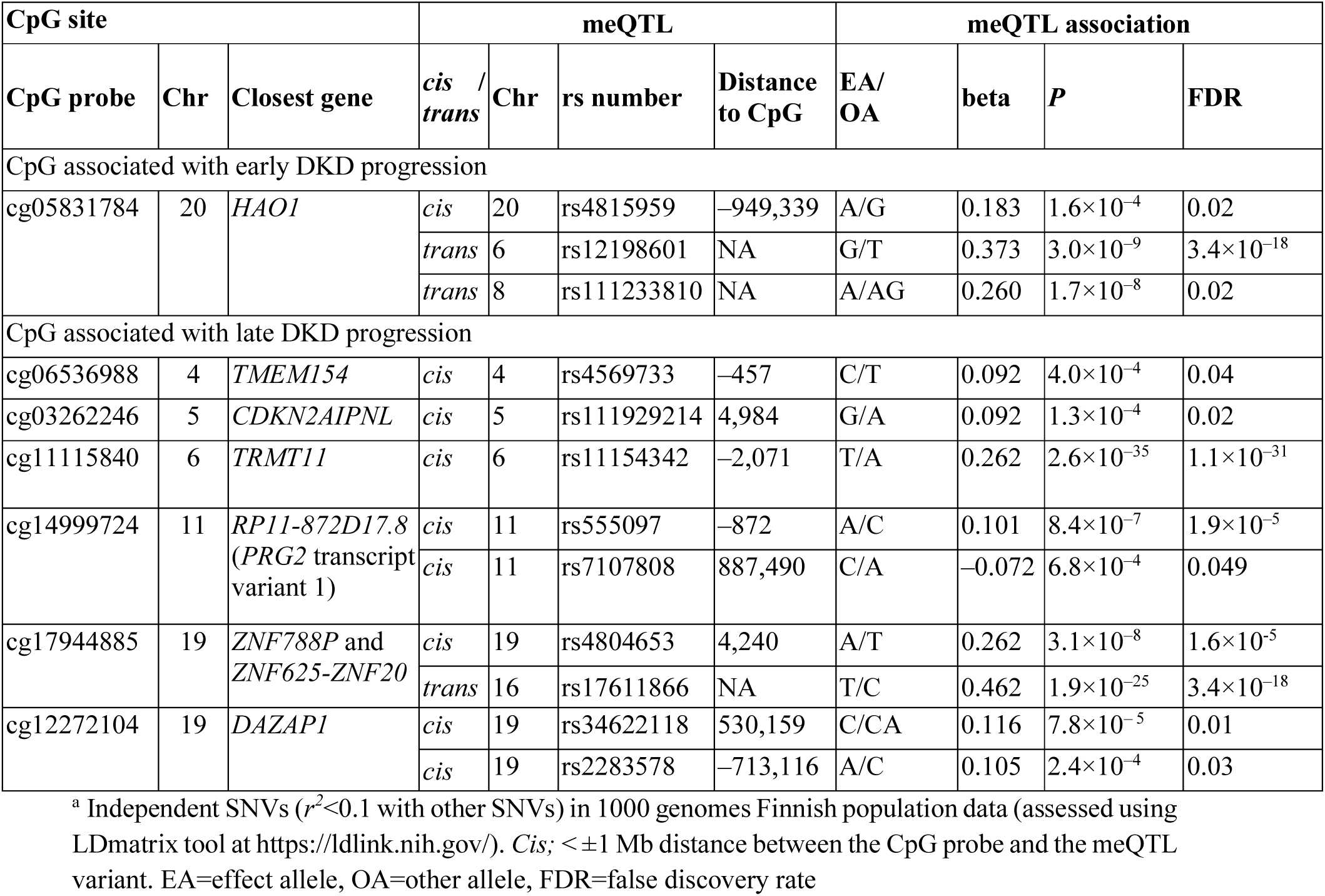
Significant independent^a^ methylation quantitative loci calculated with 765 participants from the FinnDiane study.

The 68 *trans*-meQTLs on chromosome 16 for cg17944885 were in linkage disequilibrium (*r^2^*>0.19, 1000 Genomes Finnish population data; LDlink).^49^ This locus affects (in *trans*) the expression of zinc finger genes at chr19p13.2^50^ and methylation at several loci^19^. The lead *trans*-meQTL rs17611866, a missense variant p.Val325Ala in *ZNF75A*, associates with the expression of nearby genes.^51^ Interestingly, three of the 45 CpGs regulated by rs17611866^19^ showed significant (cg17944885, chr19p13.2) or suggestive (*P*<10^−4^; cg18470038 [chr12] and cg06158227 [chr15]) association with late DKD progression in our EWAS (**Figure 5**). Furthermore, cg06158227 (chr15) was identified in an eGFR-EWAS.^13^

**Figure 5.**
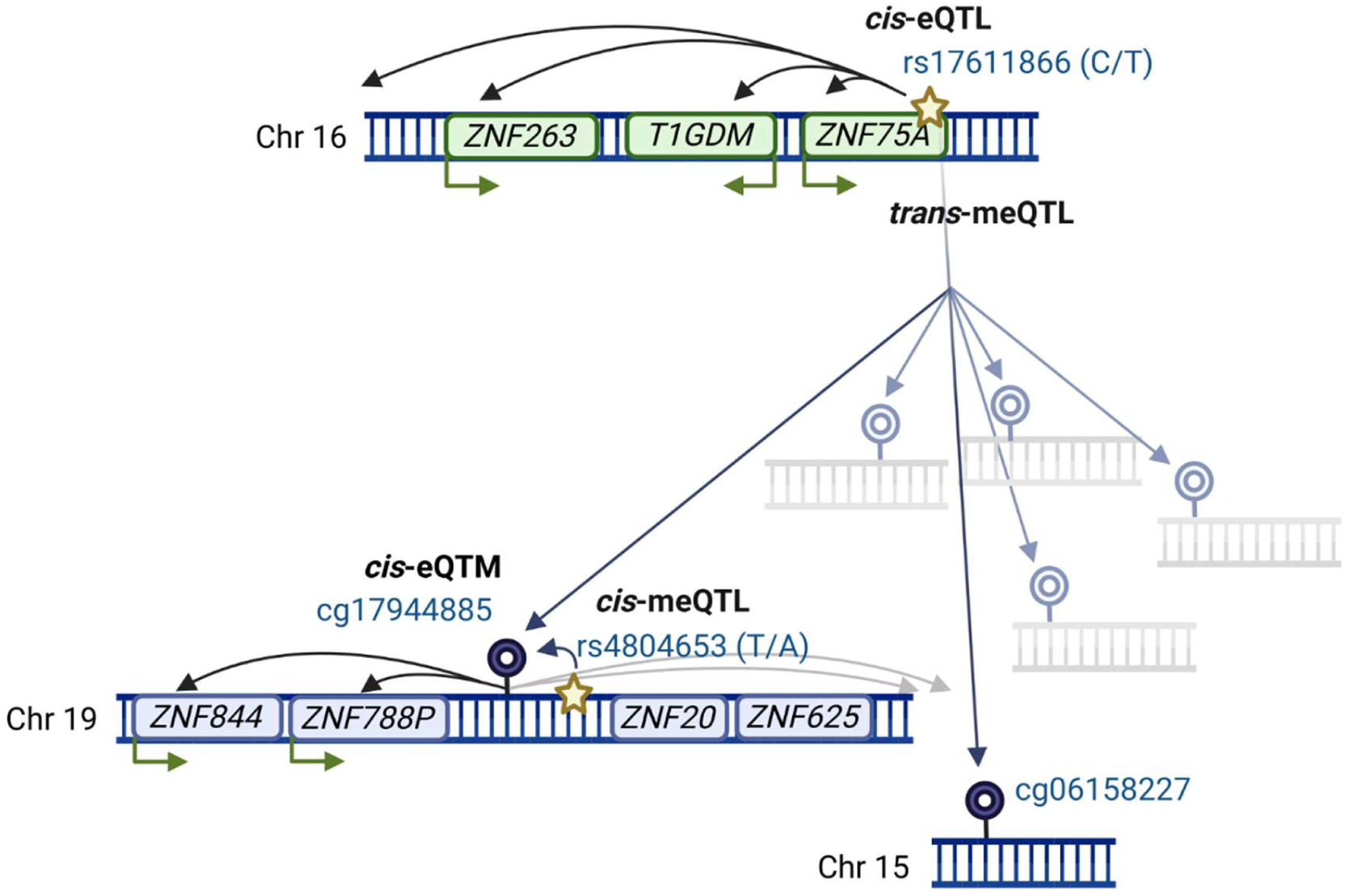
Links between methylation and gene expression of trans-meQTL locus on chromosome 16. According to Huan *et al*^19^, SNV rs17611866 correlates (in *trans*) with methylation levels of 45 CpGs, of which eGFR-associated methylation sites cg17944885 (chr19p13.2 locus, in multiple EWASs) and cg06158227^13^ are shown in the figure. CpG cg17944885 has also a close SNV rs4804653 that is associated with its methylation levels in the general population (GoDMC) data. We replicated both the *cis*- and *trans*-methylation quantitative trait loci in our diabetes cohort. Abbreviations: *cis*-eQTL=*cis* expression quantitative trait locus (SNV that affects gene expression); *cis*-meQTL=*cis* methylation quantitative trait locus; *trans*-meQTL=*trans* methylation quantitative trait locus (SNV that associates with CpG site methylation); *cis*-eQTM=*cis*-expression quantitative trait methylation (methylation site that associates with gene expression). Created in BioRender. Syreeni, A. (2024) https://BioRender.com/.

To investigate meQTL loci, we conducted phenome-wide association studies (PheWASs) in the Finnish biobank data FinnGen^52,53^ and T1D knowledge portal.^54^ Although the robust *trans*-meQTL rs17611866 in *ZNF75A* showed no significant associations, rs1447267563 near *ZNF75A* was the lead variant for “*cystic kidney disease*” and among the lead loci for “*Congenital malformations of the urinary system*”, supporting the link between this locus and kidney health. Furthermore, rs555097 (meQTL for cg14999724/*RP11-872D17.8*) associated with Cystatin C, rs12198601 (cg05831784/*HAO1*) with DKD, and rs34622118 (cg12272104/*DAZAP1*) with ESKD vs. macroalbuminuria analysis (**Supplemental Table 6**). Altogether, these associations between kidney traits and meQTLs support the importance of our top methylation sites in kidney disease.

### Gene and protein expression evidence

We investigated whether our top methylation sites were associated with gene expression. In blood cells, only cg17944885 was a significant *cis*-eQTM (**Table 4**). Remarkably, when examining data on other tissues including kidneys, 8 of 14 CpGs were significant eQTM for the closest gene (**Table 4, Supplemental Table 7**).

**Table 4.**
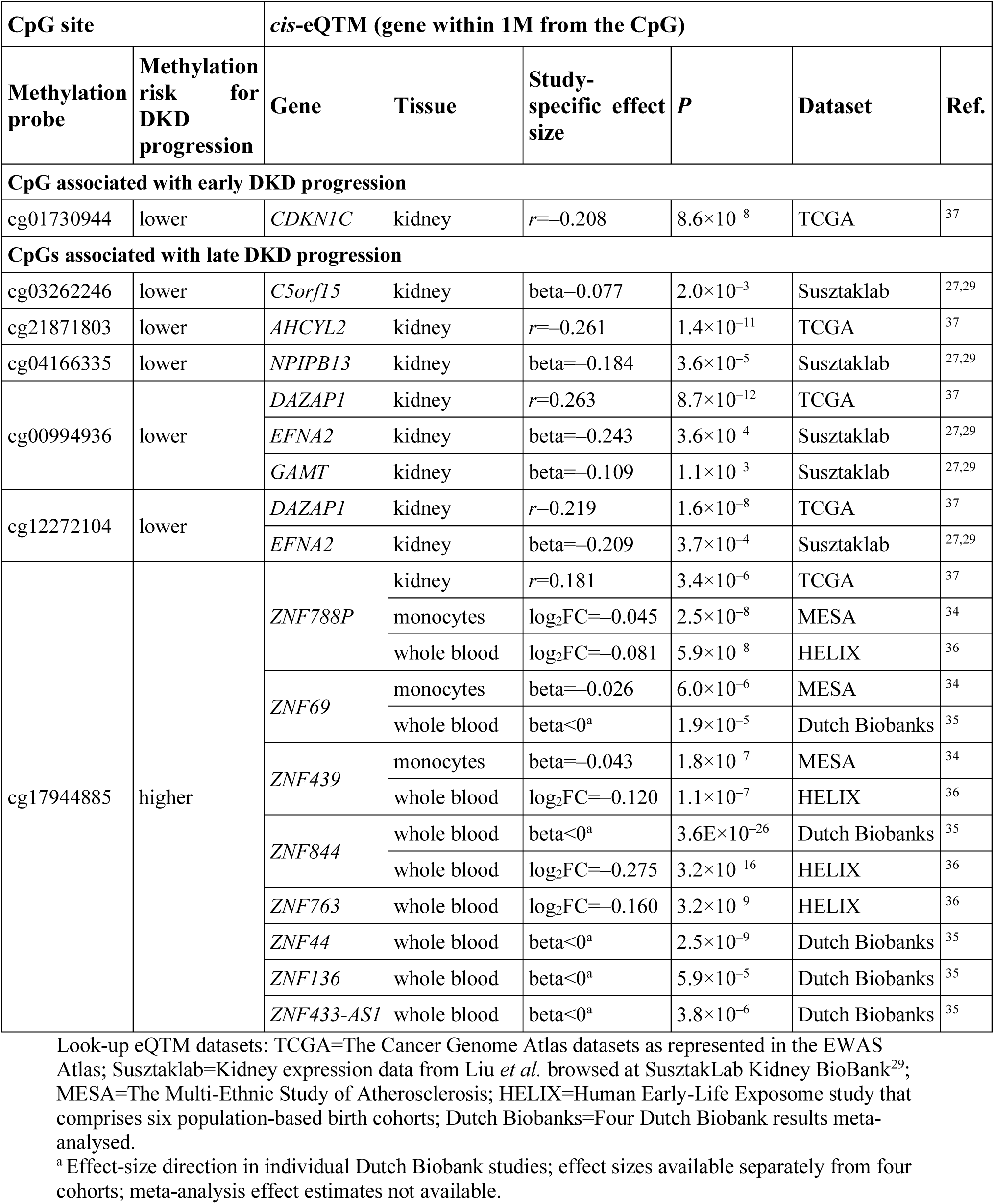
Significant *cis* expression quantitative trait methylation (*cis*-eQTM) loci in look-up analysis of 14 methylation sites for DKD progression in blood cell and kidney tissue datasets.

Our *cis*-pQTM analysis showed that cg14999724 methylation was associated with serum proteoglycan 3 levels, produced by the nearby *PRG3* gene (beta=–0.18, SE=0.04, *P*=1.7×10^−5^, **Supplemental Figure 13, Supplemental Table 8**). While *PRG3* shows limited expression in kidneys, it is over-expressed in CKD tubules^41^ and collecting duct in diabetes^46,47^ (**Supplemental Figure 14**).

We additionally studied whether the closest or eQTM-genes for the top CpGs show altered expression in kidney disease. Notably, for 13 of 14 CpGs, the related gene was differentially expressed in CKD/DKD (*P*<1.5×10^−3^) or associated with eGFR in human kidneys (**Supplemental Table 9**). For example, *CDKN1C* (near cg01730944) showed lower expression in DKD in glomeruli^38^ (FC=−4.95, **Figure 3E**) and tubules^40^ (FC=−1.55). Additionally, *AHCYL2* (near cg21871803) expression in glomeruli and tubules correlated with kidney function (*r*=0.34).^40^ For cg17944885 (chr19p13.2), four zinc finger eQTM-genes were nominally or significantly (*ZNF136*) upregulated in CKD tubules.^41^

In whole kidney samples^44^, 13 related genes were differentially expressed in advanced *vs.* early DKD, implying true biological differences related to the disease stage and justifying separate analyses like ours (**Supplemental Table 10**).

### Open chromatin and TFs

Early DKD progression-associated cg05831784 (*HAO1*), cg01730944 (*CDKN1C,* **Figure 3C**), and cg06334496 (*TMEM70*) located at open chromatin peaks^27^ in kidneys, thus, at potential regulatory regions or actively transcribed DNA. The late DKD progression-associated loci located outside of open chromatin.

Furthermore, CpGs associated with early DKD progression overlapped with several TF motifs^30^ (**Supplemental Table 11**). For example, cg01730944 (*CDKN1C*) overlapped with EGR1 that is upregulated in hyperglycemia^55^, exacerbates mesangial cell proliferation^55^, and contributes to tubular fibrosis in diabetes^56^. Taken together, snATAC-seq and TF analyses suggest that genomic regions at the novel early DKD progression -associated CpGs might have functional implications and, thus, potential relevance regarding disease progression.

### Enrichment analysis

Genes related to CpGs with EWAS *P*<1×10^−4^ were not enriched in GO terms or KEGG pathways at FDR<0.05 (**Supplemental Figures 15** and **16**). In trait enrichment analysis, early DKD progression-associated CpGs were enriched in “*exposure on glucocorticoids*” EWAS results^57^ (OR=4.5, *P*=1.3×10^−4^). Notably, glucocorticoids are anti-inflammatory medications used to improve kidney function in non-diabetic kidney disease. For late DKD progression, “*estimated glomerular filtration rate*” and “*kidney disease*” were among the enriched traits, demonstrating the consistency of our prospective EWAS with previous studies (**Supplemental Figure 17)**.

## DISCUSSION

We and others have reported cross-sectional associations between DNA methylation and DKD or eGFR and have explored the potential of CpG methylation to predict ESKD.^7,21^ To our knowledge, this is the first EWAS on early progression of DKD in T1D, and the largest study to investigate CpGs associated with late progression of DKD to ESKD. We identified four novel loci for early DKD progression, including the podocyte-specific *CDKN1C* locus. For late DKD progression, we discovered nine loci — including two previously reported and four novel sites with significant replication support from EWASs on eGFR, eGFR slope, or risk of ESKD.

Methylation levels at the CpGs associated with early DKD progression were not associated with eGFR in our data, nor in other EWASs on eGFR. Furthermore, similar early DKD progression EWAS datasets are lacking, complicating efforts to find supportive evidence. Interestingly, CpGs at *CDKN1C*, the closest gene to cg01730944, were differentially methylated in individuals with diabetes on hemodialysis, in a study of 27,000 methylation sites in saliva samples.^58^

*CDKN1C* is expressed almost exclusively in podocytes^46^, the key cell type for glomerular filtration. The Cancer Genome Atlas kidney expression data^37^ suggest that lower methylation at cg01730944 (risk of DKD progression) may be linked to *higher CDKN1C* expression; however, human DKD kidney datasets consistently showed *lower CDKN1C* expression. Thus, further eQTM evidence for cg01730944 is needed. Nevertheless, proximity to the TSS and overlap with several putative TF motifs suggest that cg01730944 methylation might regulate transcription. Notably, *EGR1,* a TF with a DNA-binding motif overlapping cg01730944, was upregulated in podocytes in individuals with diabetic nephropathy and preserved eGFR.^46^ Further, JASPAR TF data show that podocyte-specific KLF15 binds at the cg01730944 location. Importantly, *KLF15* overexpression in proteinuric mice was concomitant with upregulation of *Cdkn1c* and improved kidney health.^59^ Thus, previous research suggests that cg01730944 locus is important for kidney health, although more direct evidence is still needed. Notably, *CDKN1C* expression is regulated by the imprinting control region ICR2 such that *CDKN1C* is expressed mainly from the maternal allele, whereby loss of methylation at ICR2 decreases the expression.^60,61^

The late DKD progression-associated cg17944885 (chr19p13.2) and cg00994936 (*DAZAP1*) are known eGFR loci, first identified by Chu *et al.*^13^ Here, we identified seven novel CpGs for ESKD risk in individuals with severe albuminuria. These sites were also associated with eGFR in our study, and CpGs at *AHCYL2*, *TAOK2*, *CDKN2AIPNL,* and *RP11-872D17.8* also in other eGFR EWASs.^13–15^ Importantly, the association between cg14999724 (*RP11-872D17.8*) and ESKD risk was replicated in another prospective EWAS.^21^ We additionally identified a novel *cis*-meQTL rs555097 for cg14999724 and showed that a decrease in cg14999724 methylation (risk of ESKD) was associated with increase in serum PRG3 protein levels, in our data. However, we only found a trend in PRG3 levels between the meQTL genotypes, thus, at this site, no direct link can yet be drawn from the genetic variant, through methylation, to protein levels. While proteoglycans are components of the endothelial cell glycocalyx, a protective barrier often disrupted in diabetes-related microvascular complications^62^, proteoglycan PRG3 is primarily expressed in the bone marrow. Nevertheless, *PRG3* is overexpressed in kidney tubules in CKD. Thus, further research is needed to study its role in DKD.

The novel methylation site cg21871803 for ESKD risk, with supporting evidence from eGFR EWASs, is in *AHCYL2* (**Supplemental Figure 18**). AHCYL2 hydrolyzes S-adenosyl-L-homocysteine into adenosine and L-homocysteine, a uremic toxin increased in CKD.^63^ Kidney gene expression data suggests that lower cg21871803 methylation (risk of ESKD) correlates with *higher AHCYL2* expression. However, human kidney data are inconclusive: *AHCYL2* was upregulated in CKD^42^ but downregulated in advanced DKD.^44^

We noticed a strong genetic influence on some methylation sites: We identified ten novel meQTLs and replicated *cis*- and *trans*-meQTLs for cg17944885. Interestingly, despite a high heritability of *h^2^*=0.4^19^ and robust meQTLs, *i.e*., high genetic influence, cg17944885 methylation does not seem to be causal for DKD.^7^ Thus, kidney function decline might trigger systemic perturbations that, possibly through meQTL loci, lead to cg17944885 hypermethylation. Indeed, *trans*-meQTL locus genes *ZNF75A* and *ZNF200* were downregulated in DKD (Nephroseq) and *ZNF75A* was under-expressed in individuals on hemodialysis due to CKD.^64^

The cg17944885 locus (chr19p13.2) zinc finger TFs participate in silencing of endogenous retroviral sequences^65^, transposable elements whose elevated levels exacerbate kidney disease progression.^66^ Notably, chr19p13.2 locus genes are mostly upregulated in CKD tubules (Nephroseq), although cg17944885 hypermethylation (risk of ESRD) associate with lower expression in blood cells. Moreover, cg17944885 methylation appears dynamic: our longitudinal data showed a nominal increase in methylation in individuals with progressing DKD during follow-up. Further, blood-derived hypermethylation at cg17944885 reversed to normal after kidney transplantation.^67^ As the most replicated methylation site for kidney function, blood-derived methylation at cg17944885 is a potential general biomarker that, along with clinical factors and baseline eGFR, significantly improved the survival model for ESKD when combined with other methylation sites. Indeed, methylation risk scores for disease prediction are emerging.^68,69^

Our prospective data are unique, but the study setting has its limitations. Individuals in the early DKD progression cohort had normal AER and good to moderate kidney function despite long-lasting diabetes. Notably, most individuals in this cohort were included in our cross-sectional EWAS^7^, and unlikely included individuals with rapid DKD progression after diabetes onset. Moreover, we did not evaluate prospective albuminuria. Therefore, some individuals with persisting eGFR>60 mL/min/1.73m^2^ may have developed albuminuria during follow-up potentially diluting our associations based on eGFR decline. Additionally, eGFR declines with aging, which we accounted for by adjusting the analysis for baseline age. Despite these limitations, we identified methylation sites near relevant genes, associated with future progression to DKD.

To conclude, our two prospective EWASs on the progression of DKD in T1D identified novel methylation sites for kidney disease progression and highlighted again cg17944885 as a lead locus in kidney disease. Our findings support the role of a podocyte marker *CDKN1C* for the initiation of DKD and provide further evidence that DNA methylation can be used as a dynamic marker to improve prediction of early and late progression of DKD.

## Disclosures

P.-H. G. has received investigator-initiated research grants from Eli Lilly and Roche, is an advisory board member for AbbVie, Astellas, AstraZeneca, Bayer, Boehringer Ingelheim, Eli Lilly, Janssen, Medscape, Merck Sharp & Dohme, Mundipharma, Nestlé, Novartis, Novo Nordisk, and Sanofi; and has received lecture fees from Astellas, AstraZeneca, Bayer, Boehringer Ingelheim, Eli Lilly, Elo Water, Genzyme, Merck Sharp & Dohme, Medscape, Novartis, Novo Nordisk, PeerVoice, Sanofi, and Sciarc. S.M. has received lecture honoraria from Encore Medical Education.

## Funding

Research reported in this publication was supported by the National Institute of Diabetes And Digestive and Kidney Diseases of the National Institutes of Health under Award Numbers R01DK105154, R01DK132299, R01DK065073, and R01DK081705. The content is solely the responsibility of the authors and does not necessarily represent the official views of the National Institutes of Health. The FinnDiane study was supported by grants from Folkhälsan Research Foundation, Wilhelm and Else Stockmann Foundation, Liv och Hälsa Society, State funding for university-level health research by Helsinki University Hospital (TYH2023403), Sigrid Jusélius Foundation (220027), Novo Nordisk Foundation (NNF23OC0082732), Academy of Finland (316664), the Finnish Diabetes Research Foundation, and Medical Society of Finland (Finska Läkaresällskapet). L.J.S. was the recipient of a Northern Ireland Kidney Research Fund Fellowship. L.J.S, C.H and A.J.M were supported by an award from HSC R&D Division STL/5569/19 and UK Research and Innovation Medical Research Council MC_PC_20026. For Joslin Kidney Study, support was also obtained from the Wanek family project for the cure of Type 1 diabetes at the City of Hope Beckman Research Institute.

## Acknowledgements

We are indebted to the late Carol Forsblom (1964–2022), for his considerable contribution to the FinnDiane study. We thank all FinnDiane participants and study nurses and physicians at the study centers (**Supplemental Figure 19**). The participants and researchers of all look-up cohorts and datasets utilised in this study are greatly appreciated. We also want to acknowledge the participants and investigators of the FinnGen study. Members of the GENIE Consortium are listed in **Supplemental Figure 20**.

## Author Contributions

A.S. designed the study, analysed the FinnDiane EWAS data, performed the downstream analyses, and drafted the manuscript. E.H.D. participated in the FinnDiane methylation data collection and QC and run meQTL analyses. L.J.S. generated the methylation EPIC data for the FinnDiane and UK-ROI cohorts, and quality-controlled and analysed the EWAS data of UK-ROI and NICOLA cohorts. C.H. analysed the human kidney expression data of Fan et al and Levin et al. S.M. quality-controlled the OLINK^®^ protein data for the FinnDiane. V.H. and P.-H.G. acquired funding and phenotypic data for the FinnDiane study. Z.C. ran the EWAS in the JKS cohort and provided the replication results. R.N. contributed the EWAS data of the JKS. A.S.K. contributed to the JKS cohort data acquisition and analysis. A.P.M. contributed to the interpretation of the data and UK-ROI cohort data. A.J.M. designed the study, acquired, and analysed UK-ROI collection data and contributed to the interpretation of the results of the study. N.S. designed the study, contributed to the FinnDiane data collection and interpretation of the results of the study. Additionally, A.S., E.H.D., L.J.S., C.H., Y.G., V.H., Z.C., R.N., A.S.K., J.N.H., J.C.F., A.P.M., P.-H.G., A.J.M., and N.S. read and reviewed the manuscript draft and all authors approved the final version.

## Data Sharing Statement

The informed consent written by the participants does not allow the public sharing of the FinnDiane data analysed during the current study. Readers can propose co-operative research through the corresponding authors on reasonable request. Lookups on the supporting evidence, meQTLs, eQTMs, kidney single-cell gene expression, and DKD kidney gene expression datasets are based on published summary statistics downloadable or browsable online and access to these data sets are described in Supplemental Methods. The GWAS summary statistics of the Finnish biobank FinnGen study data freeze 10 was accessed at (https://r10.finngen.fi).

## Notes

### Author Declarations

Ethics Committee of Helsinki University Central Hospital (Helsinki, Finland) gave ethical approval of this work.

## References

1. Jansson Sigfrids F, Groop PH, Harjutsalo V. Incidence rate patterns, cumulative incidence, and time trends for moderate and severe albuminuria in individuals diagnosed with type 1 diabetes aged 0-14 years: a population-based retrospective cohort study. Lancet Diabetes Endocrinol. 2022;10(7):489–498 doi:10.1016/S2213-8587(22)00099-7

2. Salem RM, Todd JN, Sandholm N, Cole JB, Chen WM, Andrews D, et al. Genome-Wide Association Study of Diabetic Kidney Disease Highlights Biology Involved in Glomerular Basement Membrane Collagen. J Am Soc Nephrol. 2019;30(10):2000–2016 doi: 10.1681/ASN.2019030218

3. Sandholm N, Cole JB, Nair V, Sheng X, Liu H, Ahlqvist E, et al. Genome-wide meta-analysis and omics integration identifies novel genes associated with diabetic kidney disease. Diabetologia. 2022;65(9):1495–1509 doi:10.1007/s00125-022-05735-0

4. Sandholm N, Dahlström EH, Groop PH. Genetic and epigenetic background of diabetic kidney disease. Front Endocrinol. 2023;14:1163001 doi:10.3389/fendo.2023.1163001

5. Bell CG, Teschendorff AE, Rakyan VK, Maxwell AP, Beck S, Savage DA. Genome-wide DNA methylation analysis for diabetic nephropathy in type 1 diabetes mellitus. BMC Med Genomics. 2010;3:33 doi:10.1186/1755-8794-3-33

6. Smyth LJ, Patterson CC, Swan EJ, Maxwell AP, McKnight AJ. DNA Methylation Associated With Diabetic Kidney Disease in Blood-Derived DNA. Front Cell Dev Biol. 2020;8:561907 doi:10.3389/fcell.2020.561907

7. Smyth LJ, Dahlström EH, Syreeni A, Kerr K, Kilner J, Doyle R, et al. Epigenome-wide meta-analysis identifies DNA methylation biomarkers associated with diabetic kidney disease. Nat Commun. 2022;13(1):7891 doi:10.1038/s41467-022-34963-6

8. Khurana I, Kaipananickal H, Maxwell S, Birkelund S, Syreeni A, Forsblom C, et al. Reduced methylation correlates with diabetic nephropathy risk in type 1 diabetes. J Clin Invest. 2023;133(4):e160959 doi:10.1172/JCI160959

9. Smyth LJ, Kilner J, Nair V, Liu H, Brennan E, Kerr K, et al. Assessment of differentially methylated loci in individuals with end-stage kidney disease attributed to diabetic kidney disease: an exploratory study. Clin Epigenetics. 2021;13(1):99 doi:10.1186/s13148-021-01081-x

10. Sheng X, Qiu C, Liu H, Gluck C, Hsu JY, He J, et al. Systematic integrated analysis of genetic and epigenetic variation in diabetic kidney disease. Proc Natl Acad Sci U S A. 2020;117(46):29013–29024 doi:10.1073/pnas.2005905117

11. Li KY, Tam CHT, Liu H, Day S, Lim CKP, So WY, et al. DNA methylation markers for kidney function and progression of diabetic kidney disease. Nat Commun. 2023 14(1):2543 doi:10.1038/s41467-023-37837-7

12. Qiu C, Hanson RL, Fufaa G, Kobes S, Gluck C, Huang J, et al. Cytosine methylation predicts renal function decline in American Indians. Kidney Int. 2018;93(6):1417–1431 doi:10.1016/j.kint.2018.01.036

13. Chu AY, Tin A, Schlosser P, Ko YA, Qiu C, Yao C, et al. Epigenome-wide association studies identify DNA methylation associated with kidney function. Nat Commun. 2017;8(1):1286 doi:10.1038/s41467-017-01297-7

14. Schlosser P, Tin A, Matias-Garcia PR, Thio CHL, Joehanes R, Liu H, et al. Meta-analyses identify DNA methylation associated with kidney function and damage. Nat Commun. 2021;12(1):7174 doi:10.1038/s41467-021-27234-3

15. Breeze CE, Batorsky A, Lee MK, Szeto MD, Xu X, McCartney DL, et al. Epigenome-wide association study of kidney function identifies trans-ethnic and ethnic-specific loci. Genome Med. 2021;13(1):74 doi:10.1186/s13073-021-00877-z

16. Chen Z, Miao F, Paterson AD, Lachin JM, Zhang L, Schones DE, et al. Epigenomic profiling reveals an association between persistence of DNA methylation and metabolic memory in the DCCT/EDIC type 1 diabetes cohort. Proc Natl Acad Sci U S A 2016;113(21):E3002–11 doi:10.1073/pnas.1603712113

17. Chen Z, Miao F, Braffett BH, Lachin JM, Zhang L, Wu X, et al. DNA methylation mediates development of HbA1c-associated complications in type 1 diabetes. Nat Metab. 2020;2(8):744–762 doi:10.1038/s42255-020-0231-8

18. Villicaña S, Bell JT. Genetic impacts on DNA methylation: research findings and future perspectives. Genome Biol. 2021;22(1):127 doi:10.1186/s13059-021-02347-6

19. Huan T, Joehanes R, Song C, Peng F, Guo Y, Mendelson M, et al. Genome-wide identification of DNA methylation QTLs in whole blood highlights pathways for cardiovascular disease. Nat Commun. 2019;10(1):4267 doi:10.1038/s41467-019-12228-z

20. Lecamwasam A, Novakovic B, Meyer B, Ekinci EI, Dwyer KM, Saffery R. DNA methylation profiling identifies epigenetic differences between early versus late stages of diabetic chronic kidney disease. Nephrol Dial Transplant. 2021;36(11):2027–2038 doi:10.1093/ndt/gfaa226

21. Chen Z, Satake E, Pezzolesi MG, Md Dom ZI, Stucki D, Kobayashi H, et al. Integrated analysis of blood DNA methylation, genetic variants, circulating proteins, microRNAs, and kidney failure in type 1 diabetes. Sci Transl Med. 2024;16(748):eadj3385 doi:10.1126/scitranslmed.adj3385

22. Thorn LM, Forsblom C, Wadén J, Saraheimo M, Tolonen N, Hietala K, et al. Metabolic syndrome as a risk factor for cardiovascular disease, mortality, and progression of diabetic nephropathy in type 1 diabetes. Diabetes Care. 2009;32(5):950–952 doi:10.2337/dc08-2022

23. Inker LA, Eneanya ND, Coresh J, Tighiouart H, Wang D, Sang Y, et al. New Creatinine- and Cystatin C-Based Equations to Estimate GFR without Race. N Engl J Med. 2021;385(19):1737–1749 doi:10.1056/NEJMoa2102953

24. Edgar RD, Jones MJ, Robinson WP, Kobor MS. An empirically driven data reduction method on the human 450K methylation array to remove tissue specific non-variable CpGs. Clin Epigenetics. 2017;9:11 doi:10.1186/s13148-017-0320-z

25. Mansell G, Gorrie-Stone TJ, Bao Y, Kumari M, Schalkwyk LS, Mill J, et al. Guidance for DNA methylation studies: statistical insights from the Illumina EPIC array. BMC Genomics. 2019;20(1):366 doi:10.1186/s12864-019-5761-7

26. Sheng X, Guan Y, Ma Z, Wu J, Liu H, Qiu C, et al. Mapping the genetic architecture of human traits to cell types in the kidney identifies mechanisms of disease and potential treatments. Nat Genet. 2021 Sep;53(9):1322–1333 doi:10.1038/s41588-021-00909-9

27. Liu H, Doke T, Guo D, Sheng X, Ma Z, Park J, et al. Epigenomic and transcriptomic analyses define core cell types, genes and targetable mechanisms for kidney disease. Nat Genet. 2022;54(7):950–962 doi:10.1038/s41588-022-01097-w

28. Yan Y, Liu H, Abedini A, Sheng X, Palmer M, Li H, Susztak K. Unraveling the epigenetic code: human kidney DNA methylation and chromatin dynamics in renal disease development. Nat Commun. 2024;15(1):873 doi:10.1038/s41467-024-45295-y

29. Susztaklab Kidney BioBank. Available from: https://susztaklab.com/

30. eFORGE-TF. Available from: https://eforge-tf.altiusinstitute.org/

31. University of California Santa Cruz (UCSC) Genome browser. Available from: https://genome-euro.ucsc.edu/

32. Min JL, Hemani G, Hannon E, Dekkers KF, Castillo-Fernandez J, Luijk R, et al. Genomic and phenotypic insights from an atlas of genetic effects on DNA methylation. Nat Genet. 2021;53(9):1311–1321 doi:10.1038/s41588-021-00923-x

33. Genetics of DNA Methylation Consortium. Available from: http://mqtldb.godmc.org.uk/

34. Kennedy EM, Goehring GN, Nichols MH, Robins C, Mehta D, Klengel T, et al. An integrated -omics analysis of the epigenetic landscape of gene expression in human blood cells. BMC Genomics. 2018;19(1):476 doi:10.1186/s12864-018-4842-3

35. Bonder MJ, Luijk R, Zhernakova DV, Moed M, Deelen P, Vermaat M, et al. Disease variants alter transcription factor levels and methylation of their binding sites. Nat Genet. 2017;49(1):131–138 doi:10.1038/ng.3721

36. Ruiz-Arenas C, Hernandez-Ferrer C, Vives-Usano M, Marí S, Quintela I, Mason D, et al. Identification of autosomal cis expression quantitative trait methylation (cis eQTMs) in children’s blood. Suderman M, Cheah KSE, Suderman M, editors. eLife. 2022;11:e65310 doi:10.7554/eLife.65310

37. EWAS Atlas. Available from: https://ngdc.cncb.ac.cn/ewas/atlas

38. Woroniecka KI, Park ASD, Mohtat D, Thomas DB, Pullman JM, Susztak K. Transcriptome analysis of human diabetic kidney disease. Diabetes. 2011;60(9):2354– 2369 doi:10.2337/db10-1181

39. Schmid H, Boucherot A, Yasuda Y, Henger A, Brunner B, Eichinger F, et al. Modular activation of nuclear factor-kappaB transcriptional programs in human diabetic nephropathy. Diabetes. 2006;55(11):2993–3003 doi:10.2337/db06-0477

40. Ju W, Greene CS, Eichinger F, Nair V, Hodgin JB, Bitzer M, et al. Defining cell-type specificity at the transcriptional level in human disease. Genome Res. 2013;23(11):1862– 1873 doi:10.1101/gr.155697.113

41. Nakagawa S, Nishihara K, Miyata H, Shinke H, Tomita E, Kajiwara M, et al. Molecular Markers of Tubulointerstitial Fibrosis and Tubular Cell Damage in Patients with Chronic Kidney Disease. PloS One. 2015;10(8):e0136994 doi:10.1371/journal.pone.0136994

42. Nephroseq v.5. Available from: http://www.nephroseq.org

43. Levin A, Reznichenko A, Witasp A, Liu P, Greasley PJ, Sorrentino A, et al. Novel insights into the disease transcriptome of human diabetic glomeruli and tubulointerstitium. Nephrol Dial Transplant. 2020;35(12):2059–2072 doi:10.1093/ndt/gfaa121

44. Fan Y, Yi Z, D’Agati VD, Sun Z, Zhong F, Zhang W, et al. Comparison of Kidney Transcriptomic Profiles of Early and Advanced Diabetic Nephropathy Reveals Potential New Mechanisms for Disease Progression. Diabetes. 2019;68(12):2301–2314 doi:10.2337/db19-0204

45. Hill C, Duffy S, Kettyle LM, McGlynn L, Sandholm N, Salem RM, et al. Differential Methylation of Telomere-Related Genes Is Associated with Kidney Disease in Individuals with Type 1 Diabetes. Genes (Basel). 2023;14(5):1029 doi:10.3390/genes14051029

46. Wilson PC, Wu H, Kirita Y, Uchimura K, Ledru N, Rennke HG, et al. The single-cell transcriptomic landscape of early human diabetic nephropathy. Proc Natl Acad Sci U S A. 2019;116(39):19619–19625 doi:10.1073/pnas.1908706116

47. Kidney Interactive Transcriptomics results database. Available from: http://humphreyslab.com/SingleCell/displaycharts.php

48. EWAS Toolkit. Available from: https://ngdc.cncb.ac.cn/ewas/toolkit

49. Machiela MJ, Chanock SJ. LDlink: a web-based application for exploring population-specific haplotype structure and linking correlated alleles of possible functional variants. Bioinformatics. 2015;31(21):3555–3557 doi:10.1093/bioinformatics/btv402

50. Hore V, Viñuela A, Buil A, Knight J, McCarthy MI, Small K, et al. Tensor decomposition for multiple-tissue gene expression experiments. Nat Genet. 2016;48(9):1094–1100 doi:10.1038/ng.3624

51. GTEx Portal. Available from: https://gtexportal.org/home/, v.8 data accessed

52. FinnGen freeze10. Available from: https://results.finngen.fi

53. Kurki MI, Karjalainen J, Palta P, Sipilä TP, Kristiansson K, Donner KM, et al. FinnGen provides genetic insights from a well-phenotyped isolated population. Nature. 2023;613(7944):508–518 doi:10.1038/s41586-022-05473-8

54. Type 1 Diabetes Knowledge portal. Available from: https://t1d.hugeamp.org/

55. Wang D, Guan MP, Zheng ZJ, Li WQ, Lyv FP, Pang RY, et al. Transcription Factor Egr1 is Involved in High Glucose-Induced Proliferation and Fibrosis in Rat Glomerular Mesangial Cells. Cell Physiol Biochem. 2015;36(6):2093–2107 doi:10.1159/000430177

56. Hu F, Xue M, Li Y, Jia YJ, Zheng ZJ, Yang YL, et al. Early Growth Response 1 (Egr1) Is a Transcriptional Activator of NOX4 in Oxidative Stress of Diabetic Kidney Disease. J Diabetes Res. 2018;2018:3405695 doi:10.1155/2018/3405695

57. Braun PR, Tanaka-Sahker M, Chan AC, Jellison SS, Klisares MJ, Hing BW, et al. Genome-wide DNA methylation investigation of glucocorticoid exposure within buccal samples. Psychiatry Clin Neurosci. 2019;73(6):323–330 doi:10.1111/pcn.12835

58. Sapienza C, Lee J, Powell J, Erinle O, Yafai F, Reichert J, et al. DNA methylation profiling identifies epigenetic differences between diabetes patients with ESRD and diabetes patients without nephropathy. Epigenetics. 2011;6(1):20–28 doi:10.4161/epi.6.1.13362

59. Guo Y, Pace J, Li Z, Ma’ayan A, Wang Z, Revelo MP, et al. Podocyte-Specific Induction of Krüppel-Like Factor 15 Restores Differentiation Markers and Attenuates Kidney Injury in Proteinuric Kidney Disease. J Am Soc Nephrol. 2018;29(10):2529–2545 doi:10.1681/ASN.2018030324

60. Diaz-Meyer N, Day CD, Khatod K, Maher ER, Cooper W, Reik W, et al. Silencing of CDKN1C (p57KIP2) is associated with hypomethylation at KvDMR1 in Beckwith-Wiedemann syndrome. J Med Genet. 2003;40(11):797–801 doi:10.1136/jmg.40.11.797

61. Stampone E, Caldarelli I, Zullo A, Bencivenga D, Mancini FP, Della Ragione F, et al. Genetic and Epigenetic Control of CDKN1C Expression: Importance in Cell Commitment and Differentiation, Tissue Homeostasis and Human Diseases. Int J Mol Sci. 2018;19(4):1055 doi:10.3390/ijms19041055

62. Gamez M, Elhegni HE, Fawaz S, Ho KH, Campbell NW, Copland DA, et al. Heparanase inhibition as a systemic approach to protect the endothelial glycocalyx and prevent microvascular complications in diabetes. Cardiovasc Diabetol. 2024;23(1):50 doi:10.1186/s12933-024-02133-1

63. Chen W, Feng J, Ji P, Liu Y, Wan H, Zhang J. Association of hyperhomocysteinemia and chronic kidney disease in the general population: a systematic review and meta-analysis. BMC Nephrol. 2023;24(1):247 doi:10.1186/s12882-023-03295-y

64. Zawada AM, Rogacev KS, Müller S, Rotter B, Winter P, Fliser D, et al. Massive analysis of cDNA Ends (MACE) and miRNA expression profiling identifies proatherogenic pathways in chronic kidney disease. Epigenetics. 2014;9(1):161–172 doi:10.4161/epi.26931

65. Imbeault M, Helleboid PY, Trono D. KRAB zinc-finger proteins contribute to the evolution of gene regulatory networks. Nature. 2017;543(7646):550–554 doi:10.1038/nature21683

66. Dhillon P, Mulholland KA, Hu H, Park J, Sheng X, Abedini A, et al. Increased levels of endogenous retroviruses trigger fibroinflammation and play a role in kidney disease development. Nat Commun. 2023;14(1):559. doi:10.1038/s41467-023-36212-w

67. Smyth LJ, Kerr KR, Kilner J, McGill ÁE, Maxwell AP, McKnight AJ. Longitudinal Epigenome-Wide Analysis of Kidney Transplant Recipients Pretransplant and Posttransplant. Kidney Int Rep. 2023;8(2):330–340 doi:10.1016/j.ekir.2022.11.001

68. Cheng Y, Gadd DA, Gieger C, Monterrubio-Gómez K, Zhang Y, Berta I, et al. Development and validation of DNA methylation scores in two European cohorts augment 10-year risk prediction of type 2 diabetes. Nat Aging. 2023;3(4):450–458 doi:10.1038/s43587-023-00391-4

69. Thompson M, Hill BL, Rakocz N, Chiang JN, Geschwind D, Sankararaman S, et al. Methylation risk scores are associated with a collection of phenotypes within electronic health record systems. NPJ Genomic Med. 2022;7(1):50 doi:10.1038/s41525-022-00320-1

