## Supplemental Methods and Supplemental Figures for "Blood methylation biomarkers are associated with diabetic kidney disease progression in type 1 diabetes"

Supplemental Data 1: Supplemental Methods and Supplemental Figures

**Blood methylation biomarkers are associated with diabetic kidney disease progression in type 1 diabetes**

Anna Syreeni, Emma H. Dahlström, Laura J. Smyth, Claire Hill, Stefan Mutter, Yogesh Gupta, Valma Harjutsalo, Zhuo Chen, Rama Natarajan, Andrzej S. Krolewski, Joel N. Hirschhorn, Jose C. Florez, GENIE consortium, Alexander P. Maxwell, Per-Henrik Groop, Amy Jayne McKnight, Niina Sandholm on behalf of the FinnDiane Study Group

[**Supplemental Figure 15**. Gene Ontology (GO) term enrichment results of the genes related to the early and late DKD progression –associated CpGs (*P*<10^–4^).	23](#_Toc183677214)

[**Supplemental Figure 16**. KEGG pathway enrichment results of the genes related to the early and late DKD progression –associated CpGs (*P*<10^–4^).	24](#_Toc183677215)

### 1. Supplemental Methods

#### I. DNA methylation assessment and quality control

DNA methylation of bisulphite-converted DNA samples (EZ Zymo Methylation Kit (Zymo Research, USA) were analysed with BeadChips and Infinium MethylationEPIC Kit v1.0 as in the protocol. iScan machine generated the methylation intensity files (.idat). We then estimated six proportional white blood cell counts (WCCs) from the raw .idat files using the Houseman method^1^ with Bioconductor v3.10 and '*minfi*'-package’s estimateCellCounts -function. The quality control (QC) was performed jointly for all 898 sample .idat files using 'RnBeads' v.2.6.0 through R v4.0.0. The QC included normalization with bmiq, removal of cross-reactive probes, probes in sex chromosomes or those near SNVs. In QC, from the initial 898 samples and 866,895 methylation probes, one sample (greedycut *P*<0.05) and 105,357 methylation probes were removed (**Table 1**).

**Table 1** Methylation data preprocessing

| Step | Number of probes | Samples |
| --- | --- | --- |
| Numbers prior pre-processing | 866,895 | 898 |
| removal of SNV-enriched probes | 17,371 |  |
| removal of cross-reactive probes | 43,463 |  |
| removal of probes and samples with greedycut, *P*<0.05 | 27,117 | 1 |
| removal of probes with no context | 1,072 |  |
| removal of probes in sex chromosomes | 16,334 |  |
| Numbers after pre-processing | 761,538 | 897 |

For the Epigenome-wide association study (EWAS), we extracted methylation M-values using *mval* -function in *'RnBeads'.* We additionally extracted Infinium MethylationEPIC Kit control probe intensities using *qc*-function in *'RnBeads'*. We calculated principal components (PC) from all 225 non-negative control probe red (Cy5) and green (Cy3) signal intensities with *prcomp-*function of base R with default settings except normalization was set to “TRUE”. PCs 1–3 explained >90% of the variability in these control probes, and they were used as covariates in the following EWASs.

Additionally, we calculated the mean methylation level per each sample using the extracted M values. Of the known 114,204 CpGs invariable in blood-based DNA^2^ our data included 99,249 of which 86,980 were truly invariable — defined as a range of methylation beta-values <0.05 in samples in one batch, within >50% of the technical batches. Then, we calculated the intrapersonal mean methylation (mean M) of these 86,980 sites to be used as a covariate in EWAS to correct for further technical (e*.g.,* batch) related deviations.

#### II. Methylation values for the longitudinal analyses

For 52 individuals with methylation data from two time points, we converted the M values to methylation beta-values and regressed out technical and cell composition –related variability by fitting a linear model: Δbeta ~ ΔGranulocytes + ΔB-cell + ΔCD4T-cell + ΔCD8T-cell + ΔMonocytes + ΔNK-cells + ΔPC1 + ΔPC2 + ΔPC3 + ΔMeanM, where Δ values were calculated as (Value_time point2_ – Value_time point1_) / years between). Thereafter, we used the model residuals in the subsequent longitudinal analyses. We calculated similar residualised beta-values for the baseline methylation values as well, to regress out blood cell-proportion -and technical issues -related variability. We compared methylation change over time between DKD progressors and non-progressors using logistic regression and residualised methylation delta M values and for baseline methylation values (residuals). Additionally, we used linear regression to test the association between eGFR slope between the time points (dependent variable) and Δbeta values and included baseline age as a covariate.

#### III. Sensitivity analyses

*10-year-risk* *—* The assumption of proportional-hazards in Cox models may not hold in long-term follow-up. Therefore, we repeated the Cox analyses for our top CpGs in the late DKD analysis but restricted the follow-up to ten years after baseline. Due to the smaller number of events, this analysis was not applied to the early DKD progression cohort.

*Linear association with baseline eGFR —* For the significant CpGs (*P*<9.4×10^–8^), we tested the association between log2-transformed baseline eGFR and methylation using *'limma'* with the same covariates as in the Cox proportional-hazards model. This analysis was applied to the early or late DKD progression sub-cohorts and the combined cohort, where the analysis was additionally adjusted for baseline albuminuria status (normal AER / severe albuminuria).

*Competing risk analysis* *—* For the significant CpGs in the late DKD progression cohort, we used Fine and Grey regression analysis in R4.1.3 with *'survival'*-package v3.2-13 to account for the competing risk of death. The model included the same variables as were included in the corresponding Cox model, ESKD as an event, and death as a competing risk.

*Correlation with baseline variables —* To study pleiotropy, we tested the Spearman correlation between DNA methylation of the top CpGs and the baseline clinical variables including sex, age, diabetes duration, systolic and diastolic blood pressure, BMI, central obesity, current smoking status, triglycerides concentration, total cholesterol, LDL-cholesterol, and HDL-cholesterol. As methylation values, we used both M values and residuals from a model where technical variability was regressed out.

#### IV. Transcription factor analysis

We studied the overlap of the lead methylation sites with transcription factor motifs in eFORGE-TF database v2.0.^3^ We additionally sighted the JASPAR CORE 2022 transcription factor binding data through the University of California Santa Cruz (UCSC) Genome browser.^4^

#### V. Methylation quantitative trait locus (meQTL) analysis

We performed meQTL analysis to assess both *cis* (±1 Mb) and *trans* genetic effects using R package *'Matrix eQTL'* v.2.3. For this, we included a subset of 765 FinnDiane participants that were part of our previous EWAS on DKD^5^ and that had imputed and quality-controlled genotyping data^6^ available. Here, we included 6,010,201 SNVs with a minor allele frequency ≥0.05 and imputation info ≥0.80. For the meQTL analysis, we used an additive linear model adjusted for age, sex, diabetes duration, and six WCCs. For the CpGs, we used methylation M values and window of ±1 Mb (maximum distance between SNV and CpG site) to assess *cis* effects. Additionally, we searched general population meQTLs from the Genetics of DNA Methylation Consortium (GoDMC) data of 27,750 Europeans.^7,8^

#### VI. Expression quantitative trait methylation (eQTM) data lookups

We examined published eQTM (methylation *vs.* gene expression) datasets on whole blood in adults^9,10^ or children^11^, monocytes^9^, or kidney tissue^12^ and looked up the Cancer Genome Atlas (TCGA) data of multiple tissues through EWAS Atlas.^13^

#### VII. Nephroseq v.5 human kidney gene expression datasets

We studied the expression of the genes related to the top CpG sites in human kidney datasets in Nephroseq v.5 database.^14^ The following datasets were studied for disease (DKD/CKD) vs control analysis (fold-change >1.5 and *P*<0.05 reported) and correlation with eGFR or proteinuria (*P*<0.05 reported). Significance was set to the number of related genes found in Nephroseq data (*P*<0.05/34=1.47×10^−3^). Woroniecka dataset^15^ included 22 glomerular (9 with DKD) and 22 tubular (10 with DKD) kidney biopsy samples from healthy, living transplant donors. The Schmid dataset^16^ included 24 kidney tubular biopsies: 13 from individuals with DKD and 11 from healthy controls with no kidney disease or minimal change disease. The datasets from Ju and colleagues^17^ were glomerular or tubular kidney biopsies from individuals with DKD and healthy living donors or individuals with other diseases. The European Renal cDNA Bank-Kröner-Fresenius Biopsy Bank (ERCB) dataset comprised tubulointerstitial samples of ten individuals with DKD and nine healthy living donors. Finally, two Nakagawa CKD datasets consisting of tubulointerstitial kidney biopsies of 53 (discovery set, *n*=48 with CKD) and eight (validation set, five with CKD) were examined.

#### VIII. Serum protein measurements

We utilised serum samples stored at −20ᵒC in the proteomics analysis with OLINK® Explore Ht assay. The 860 samples were randomized on 10 plates and the protein expression was measured at the SciLifeLab in Uppsala, Sweden. The protein expression values were corrected for the plate control and then intensity normalized in Uppsala to remove inter- and intra-batch variation (NPX, Normalized Protein eXpression; Log2 scale). We further excluded any individual protein values exceeding 5 standard deviations from the mean value measured from that particular protein. Three samples were excluded because of assay failures, and we further excluded 14 samples as outliers as either the overall sample median or the sample interquartile range was more than 3 standard deviations away from the mean values.

Out of 843 proteomic data quality-control-passing samples, altogether 313 were overlapping with our EWAS cohorts, thus, had methylation data from the same time-point. This overlap comprised 188 individuals with normal AER for the main protein quantitative trait methylation (pQTM) analysis and 127 individuals with macroalbuminuria for look-up replication.

We successfully mapped 5,379 OLINK panel proteins to genomic co-ordinates (hg19) of the related RefSeq gene using UCSC Table browser. Additionally, genes for 29 proteins were manually mapped with their UniportIDs to Ensemble gene id and location at [www.uniprot.org](http://www.uniprot.org) (Accessed 20 May, 2024). As a result, 106 protein products of genes located within 1Mb from the top 14 methylation CpG formed 126 methylation CpG – protein pairs for the subsequent *cis* protein quantitative trait methylation (*cis*-pQTM) analysis.

#### IX. Protein quantitative trait methylation (pQTM) analysis

First, we residualised the methylation M-values to remove variability due to technical or blood cells proportions: we fitted a linear model (CpG_methylation_M ~ Granulocytes + ΔB-cell + CD4T-cell + CD8T-cell + Monocytes + NK-cells + PC1 + PC2 + PC3 + MeanM) to the data and used rank-based inverse normal transformation (INT) normalized residuals from the model in subsequent *cis*-pQTM analysis. The linear regression model in the *cis*-pQTM included protein values as an outcome, inverse normal transformed (INT) residualised methylation values (14 CpGs analysed separately) as an independent variable and sex, age, estimated glomerular filtration rate, serum storage time, and number of thawing (≤5) as covariates. Significance limit was set by the number of analysed CpG-proteins pairs (*n*=126) to 3.97×10^−4^ (0.05/126).

#### X. References

1.	Houseman EA, Accomando WP, Koestler DC, et al. DNA methylation arrays as surrogate measures of cell mixture distribution. *BMC Bioinformatics*. 2012;13:86. doi:10.1186/1471-2105-13-86

2.	Edgar RD, Jones MJ, Robinson WP, Kobor MS. An empirically driven data reduction method on the human 450K methylation array to remove tissue specific non-variable CpGs. *Clin Epigenetics*. 2017;9:11. doi:10.1186/s13148-017-0320-z

3.	eFORGE-TF. https://eforge-tf.altiusinstitute.org/

4.	University of California Santa Cruz (UCSC) Genome browser, GrCh37 (hg19). https://genome-euro.ucsc.edu/, Accessed 2023

5.	Smyth LJ, Dahlström EH, Syreeni A, et al. Epigenome-wide meta-analysis identifies DNA methylation biomarkers associated with diabetic kidney disease. *Nat Commun*. 2022;13(1):7891. doi:10.1038/s41467-022-34963-6

6.	Sandholm N, Hotakainen R, Haukka JK, et al. Whole-exome sequencing identifies novel protein-altering variants associated with serum apolipoprotein and lipid concentrations. *Genome Med*. 2022;14(1):132. doi:10.1186/s13073-022-01135-6

7.	Min JL, Hemani G, Hannon E, et al. Genomic and phenotypic insights from an atlas of genetic effects on DNA methylation. *Nat Genet*. 2021;53(9):1311-1321. doi:10.1038/s41588-021-00923-x

8.	Genetics of DNA Methylation Consortium. http://mqtldb.godmc.org.uk/

9.	Kennedy EM, Goehring GN, Nichols MH, et al. An integrated -omics analysis of the epigenetic landscape of gene expression in human blood cells. *BMC Genomics*. 2018;19(1):476. doi:10.1186/s12864-018-4842-3

10.	Bonder MJ, Luijk R, Zhernakova DV, et al. Disease variants alter transcription factor levels and methylation of their binding sites. *Nat Genet*. 2017;49(1):131-138. doi:10.1038/ng.3721

11.	Ruiz-Arenas C, Hernandez-Ferrer C, Vives-Usano M, et al. Identification of autosomal cis expression quantitative trait methylation (cis eQTMs) in children’s blood. Suderman M, Cheah KSE, Suderman M, eds. *eLife*. 2022;11:e65310. doi:10.7554/eLife.65310

12.	Liu H, Doke T, Guo D, et al. Epigenomic and transcriptomic analyses define core cell types, genes and targetable mechanisms for kidney disease. *Nat Genet*. 2022;54(7):950-962. doi:10.1038/s41588-022-01097-w

13.	EWAS Atlas. https://ngdc.cncb.ac.cn/ewas/atlas

14.	Nephroseq v.5. http://www.nephroseq.org

15.	Woroniecka KI, Park ASD, Mohtat D, Thomas DB, Pullman JM, Susztak K. Transcriptome analysis of human diabetic kidney disease. *Diabetes*. 2011;60(9):2354-2369. doi:10.2337/db10-1181

16.	Schmid H, Boucherot A, Yasuda Y, et al. Modular activation of nuclear factor-kappaB transcriptional programs in human diabetic nephropathy. *Diabetes*. 2006;55(11):2993-3003. doi:10.2337/db06-0477

17.	Ju W, Greene CS, Eichinger F, et al. Defining cell-type specificity at the transcriptional level in human disease. *Genome Res*. 2013;23(11):1862-1873. doi:10.1101/gr.155697.113

### 2. Supplemental Figures

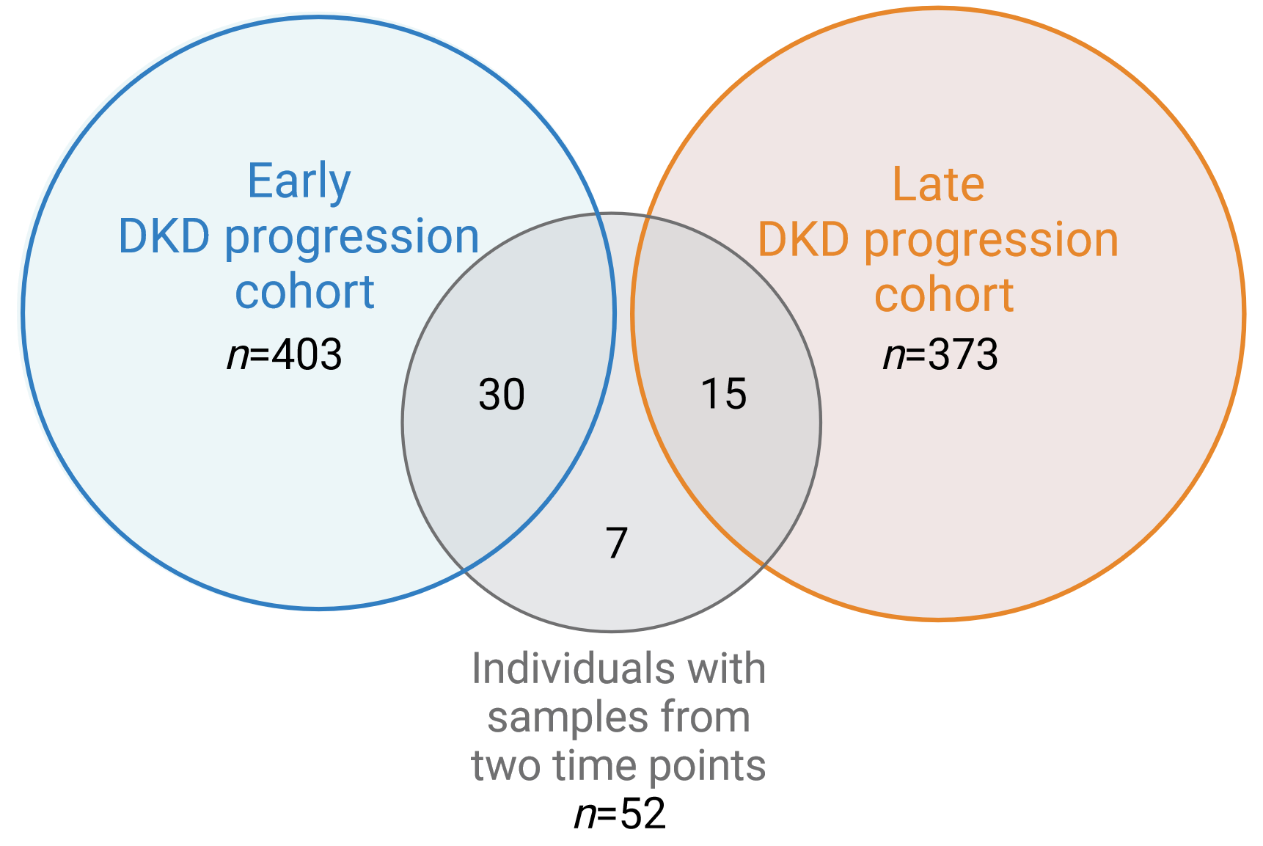

**Supplemental Figure 1**. Overlap of the individuals with longitudinal samples with the DKD progression cohorts. For the overlapping individuals, the second time point sample was always analysed in the DKD progression cohorts. Created with Biorender.com.

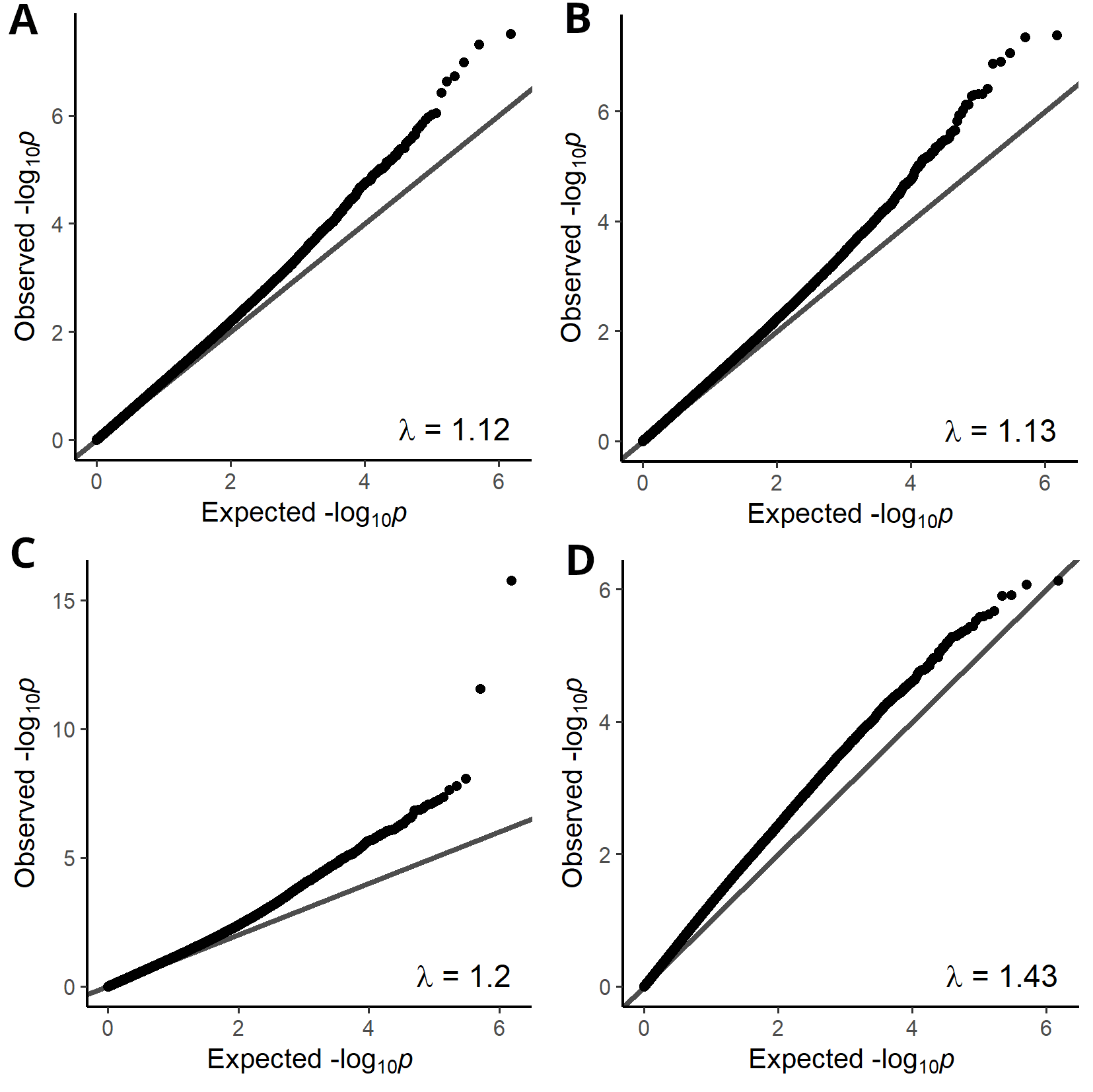

**Supplemental Figure 2**. QQ-plots of the four prospective EWASs. A) early DKD progression B) early DKD progression, adjusted for baseline eGFR, C) late DKD progression D) late DKD progression adjusted for baseline eGFR.

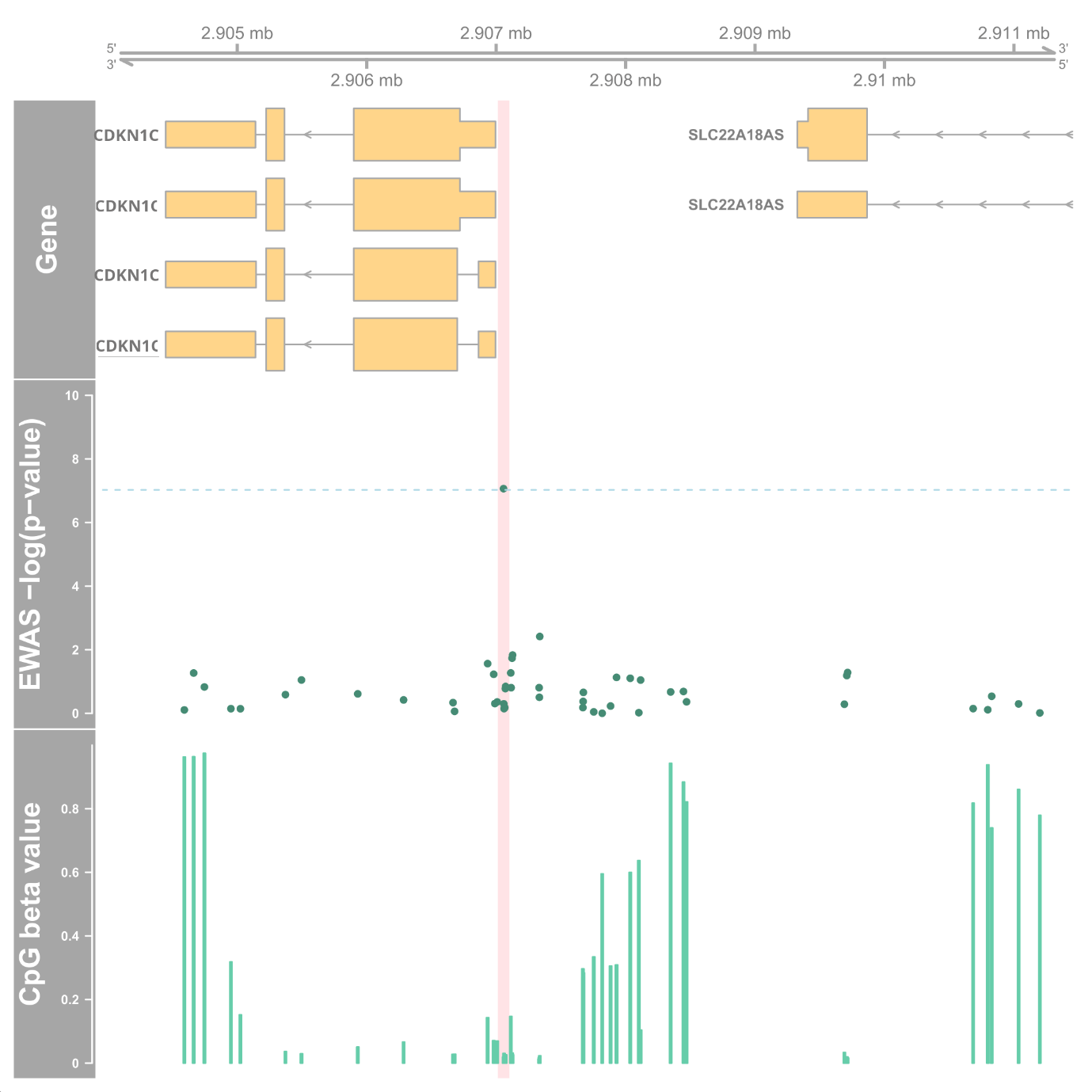

**Supplemental Figure 3.** Chromosome 11p15.5 region around cg01730944. Genes-track shows the UCSC genes and the –log(*p*-value) is for the *p*-value from the EWAS on early DKD progression in 403 individuals. Dashed blue line shows the epigenome-wide significance level at 9.4×10^–8^. The CpG beta values are mean values in the early DKD progression cohort. The light red vertical highlight shows a 100 bp region around the top CpG cg01730944. Figure generated using the *Gviz* R-package.

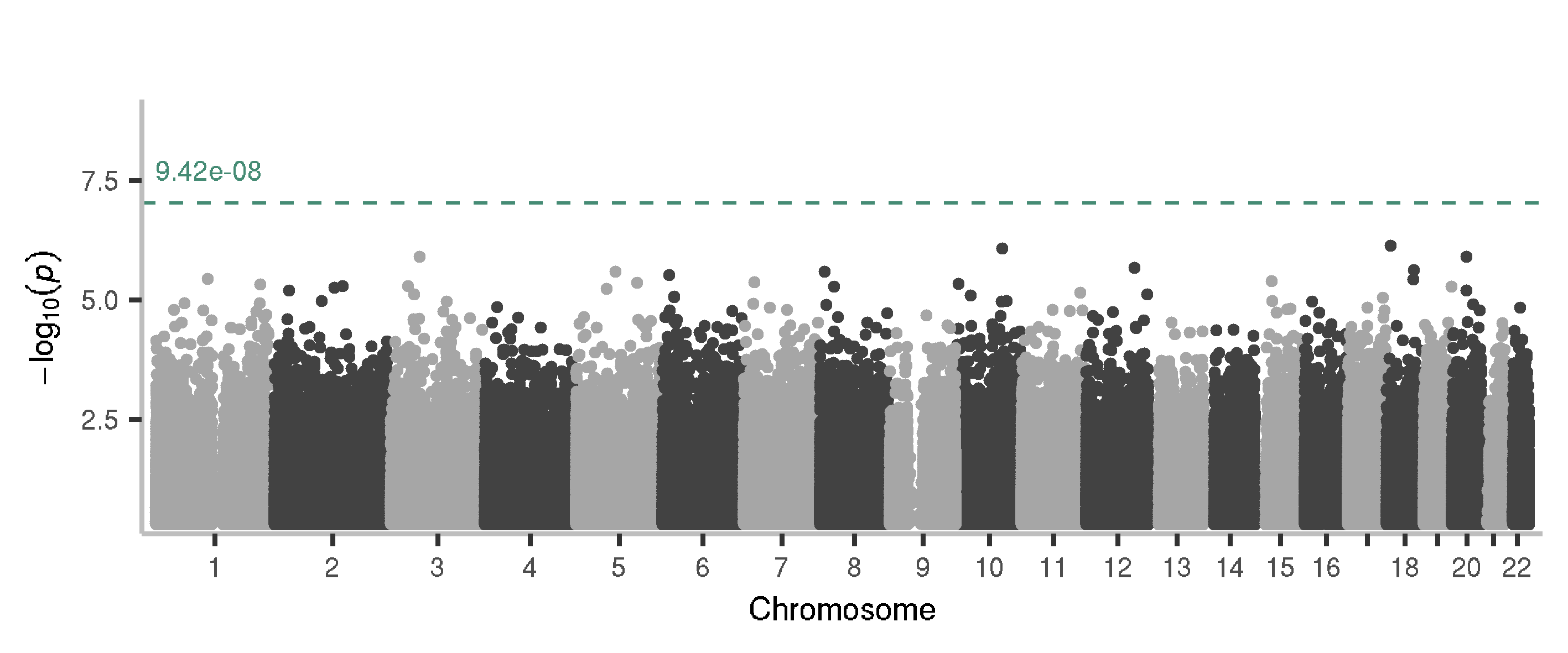

**Supplemental Figure 4.** Manhattan plot of EWAS on late progression of DKD (to end-stage kidney disease), additionally adjusted for baseline eGFR. The cohort included 373 individuals of which 206 developed ESKD. The EWAS is a Cox-proportional hazards model for ESKD event has methylation M value, age, sex, six white blood cell proportions, technical PCs 1–3, mean M from invariable sites and baseline eGFR, as covariates. Chromosomal co-ordinates are on the x-axis and y-axis shows the association significance in −log10-transformed *P*-values. As seen from the plot, no CpG site reached epigenome-wide significance.

**A**	**B**

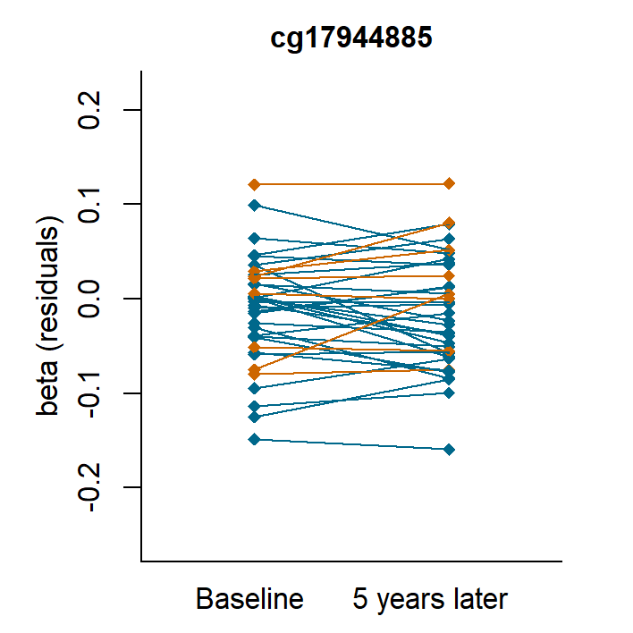

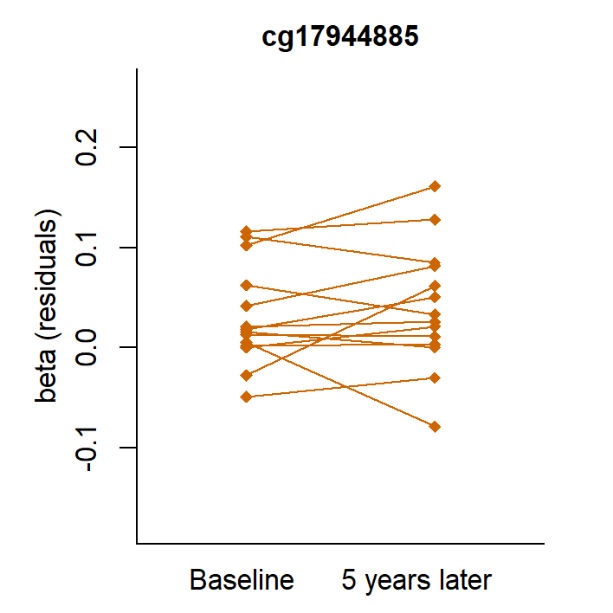

**Supplemental Figure 5**. Longitudinal change of cg17904885 methylation values (as residuals) in 52 individuals. **A)** N=38 all had normal AER at baseline and *n*=8 who progressed to severe albuminuria during follow-up are shown dark orange. The Δbeta (as residuals, technical variability regressed out) was 0.00576 in progressors and –0.00577 in non-progressors (*P=* 0.049 in logistic regression adjusted for baseline methylation level). B) N = 14 individuals with moderate albuminuria at baseline and all increased to severe albuminuria during follow-up. The baseline methylation values in both A and B figures are residuals from the following linear model: baseline_cg17944885_beta ~  Granylocyte + CD4T + CD8T + B-cell + Monocyte + NK-cell + PC1 + PC2 + PC3 + Mean M from invariable sites). The 5-year follow-up values are the sum of baseline value and residuals from a model Δcg17944885_beta ~ΔGranulocyte + ΔD4T + ΔCD8T + ΔB-cell + ΔMonocyte + ΔNK-cell + ΔPC1 + ΔPC2 + ΔPC3 + ΔMean M from invariable sites), where Δ-values were calculated as ((time point 2 value – time point 1 value)/years between) × 5 years to get a similar follow-up time for every individual.

**A**	**B**

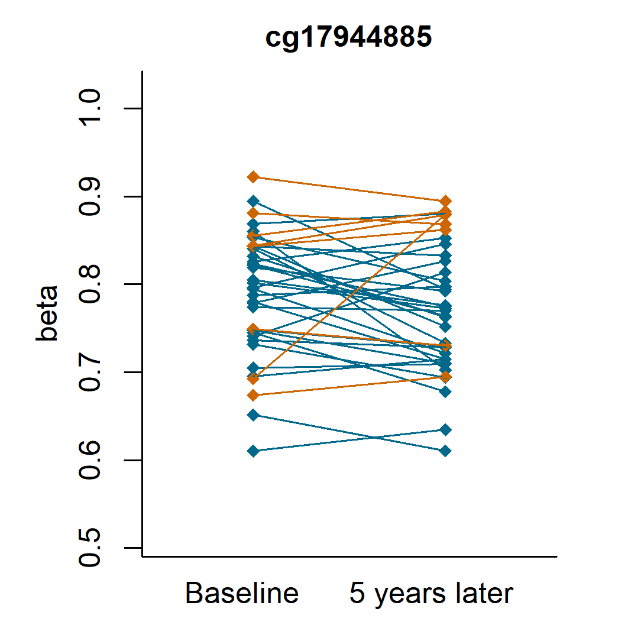

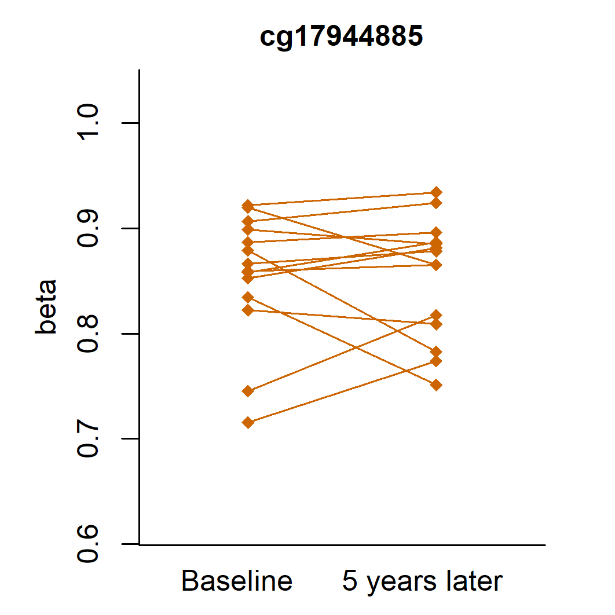

**Supplemental Figure 6**. Longitudinal change of cg17904885 methylation beta values in 52 individuals. Similar to Supplemental Figure 4. but instead of residuals, methylationn beta values were used. **A)** N=38 all had normal AER at baseline but *n*=8 progressed to severe albuminuria during the follow-up (dark orange color). The baseline beta values were 0.785 and 0.808 in non-progressors and progressors, respectively (*P*=0.53). Yearly slope of beta-value change between time points was –0.0058 in non-progressors and 0.0058 in progressors (*P*=0.05) and the 5-year time point values were 0.756 in non-progressors and 0.836 in progressors (*P*=0.02). **B)** *n*=14 individuals with moderate albuminuria at baseline and progression to severe albuminuria during follow-up. Median yearly slope of methylation beta change in this group was +0.002.

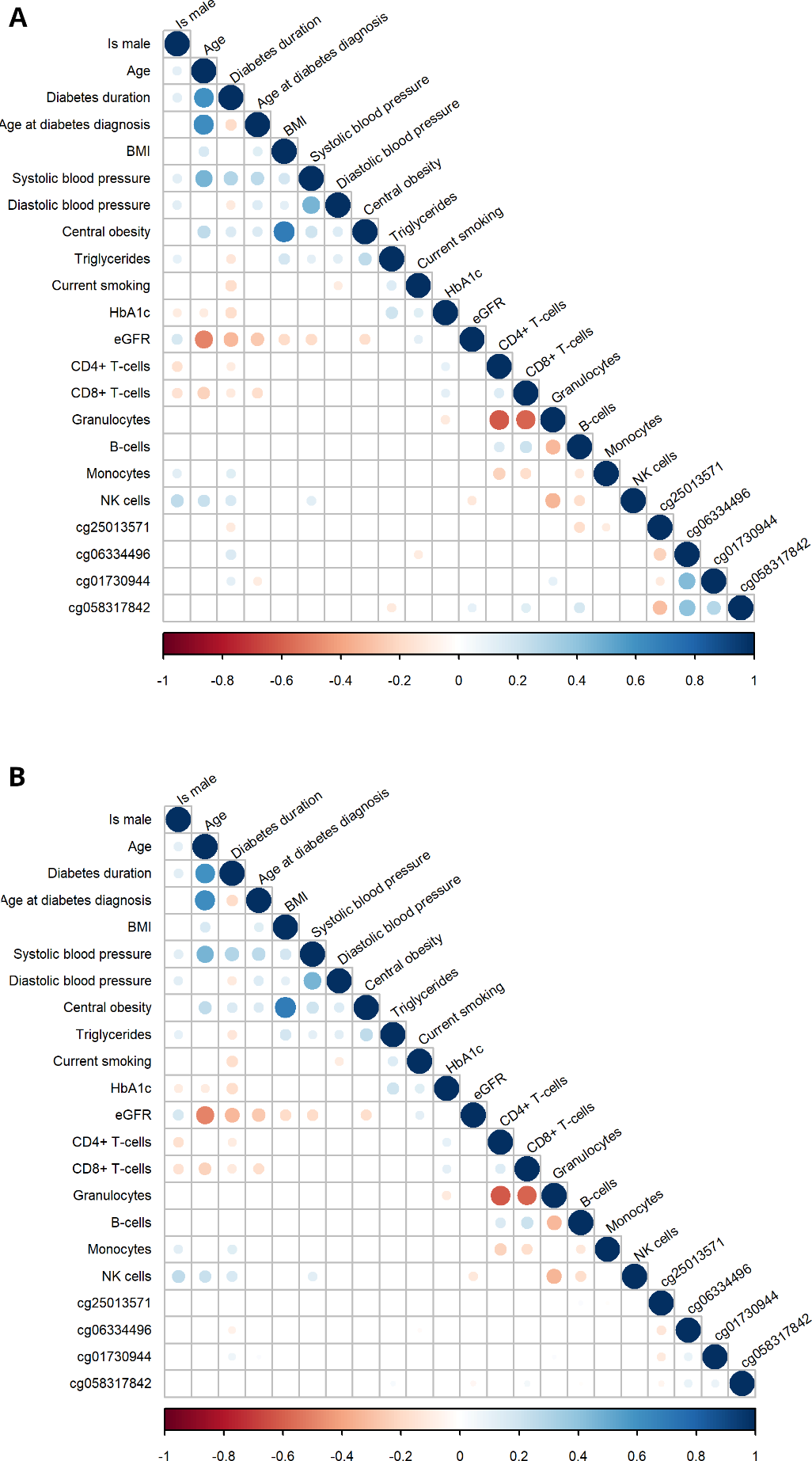

**Supplemental Figure 7.** Correlation of clinical characteristics and methylation CpGs of the early DKD progression cohort (*n*=403). A positive correlation (Spearman rho>0) is indicated in shades of blue and negative correlation in shades of red. Size of the circle refers to the strength of correlation. All correlations with *P*<0.05 are left white. **A)** M values used for the CpGs. **B)** Residualised methylation M values with technical variability regressed out (residuals from a linear model: CpG M values as dependent variable, six white blood cell counts, PCs 1–3 and mean methylation from invariable sites as variables in the model).

**A**

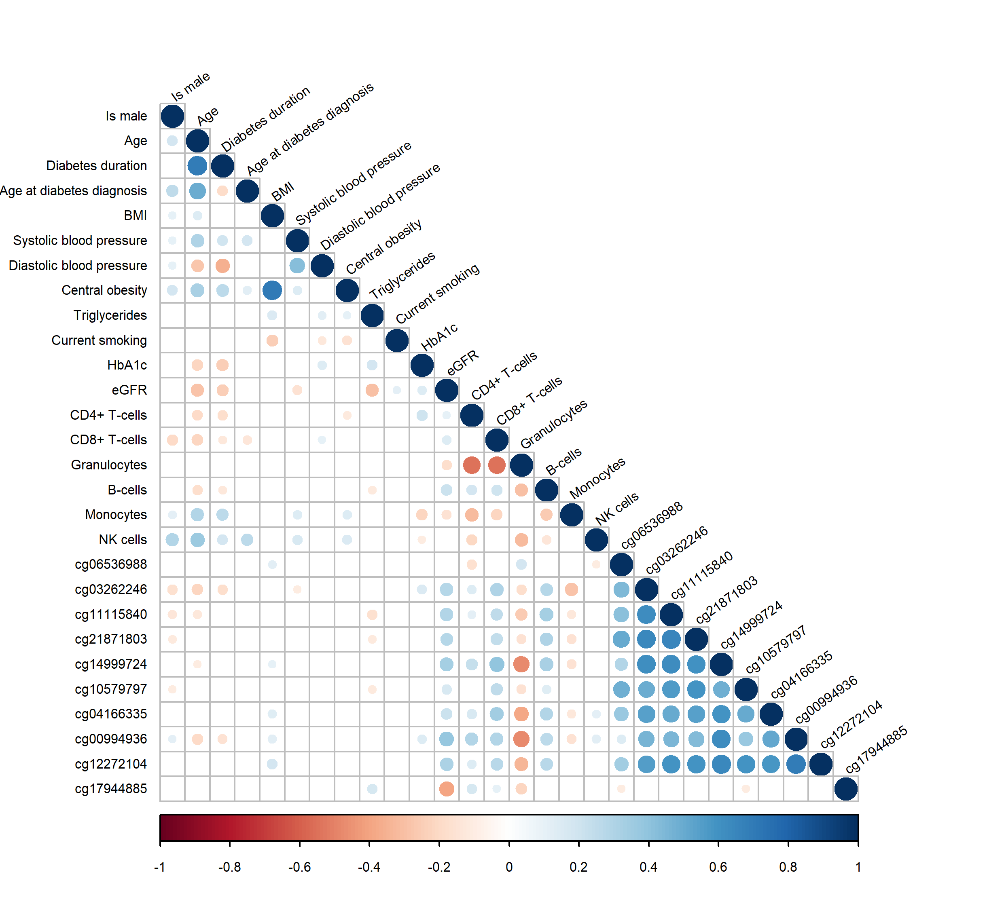

**B**

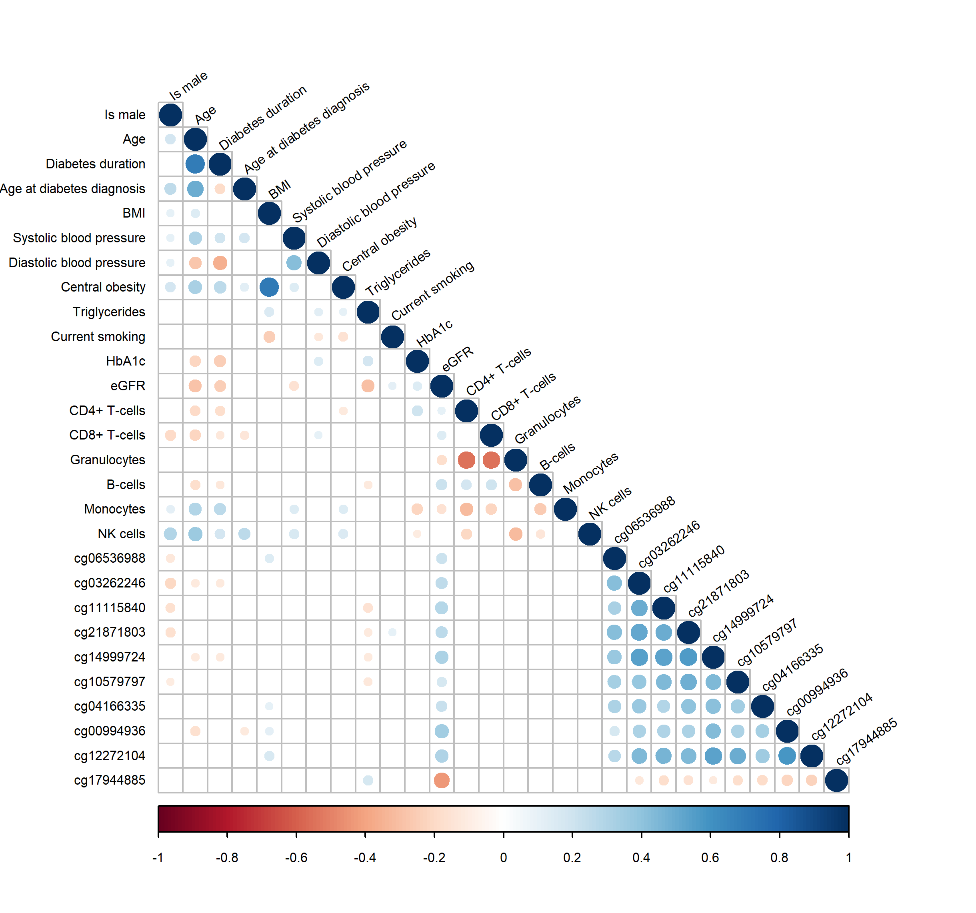

**Supplemental Figure 8.** Correlation of clinical characteristics and methylation of top CpGs in the late DKD progression cohort (*n=*373). A positive correlation (rho > 0) is indicated in shades of blue and negative correlation in shades of red. Size of the circle refers to the strength of correlation. All cells with correlations with *P*<0.05 are left white. **A)** M values used for the CpGs **B)** Technical variability regressed out from CpG methylation values (residuals from a linear model plotted: CpG M values as dependent variable, six white blood cell counts, PCs 1–3 and mean methylation from invariable sites as variables in the model).

**A**	**B**

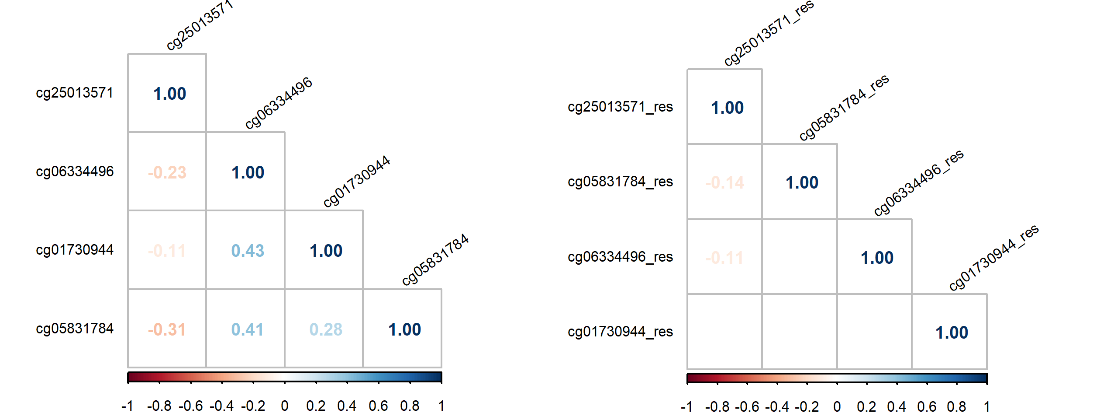

**C	D**

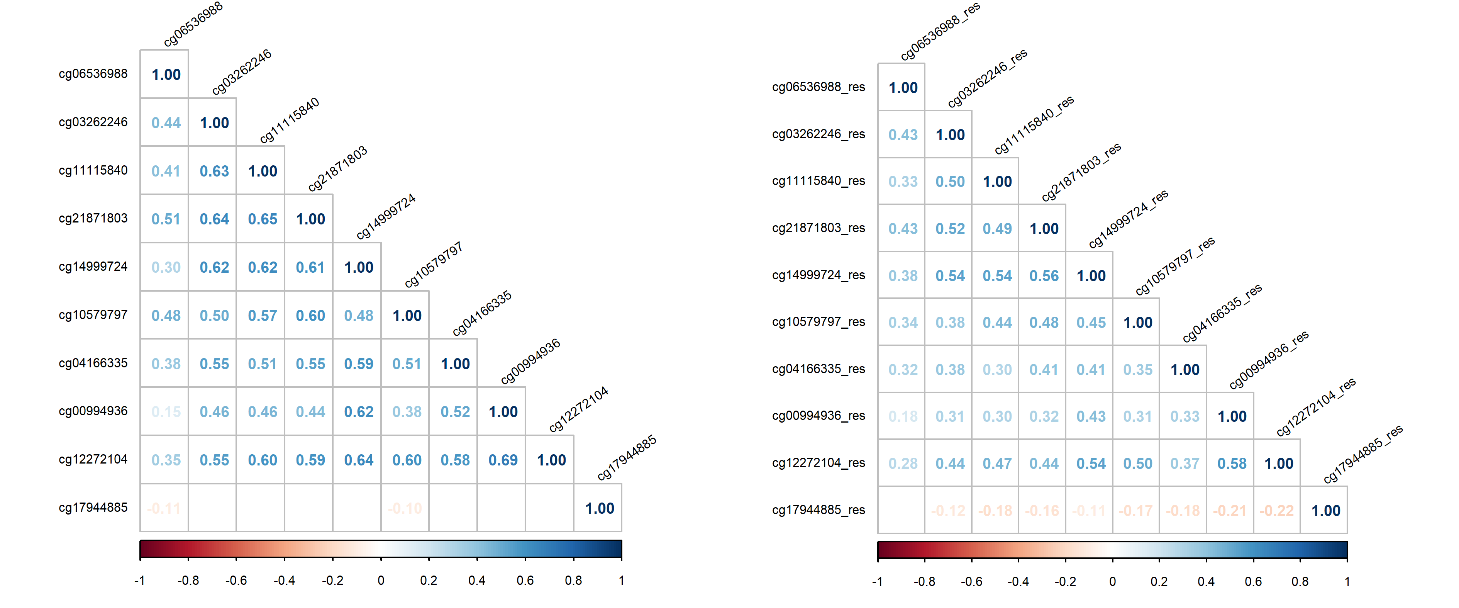

**Supplemental Figure 9.** Correlation (Spearman) of the top CpGs Panels **A** and **B** show the correlation of the four top CpGs in the early DKD progression EWAS (*n*=403 in the cohort). When A plots correlations with methylation beta-values, panel B shows the correlation of the residuals from a linear model CpG ~ PC1 + PC2 + PC3 + 6WCC + mean M from invariable sites for each CpG. Panels **C** and **D** show the correlation of the top ten CpGs in the late DKD progression EWAS (*n*=375). C panel represent correlation coefficients (rho) of the methylation beta values and D shows correlation of residualised beta values.

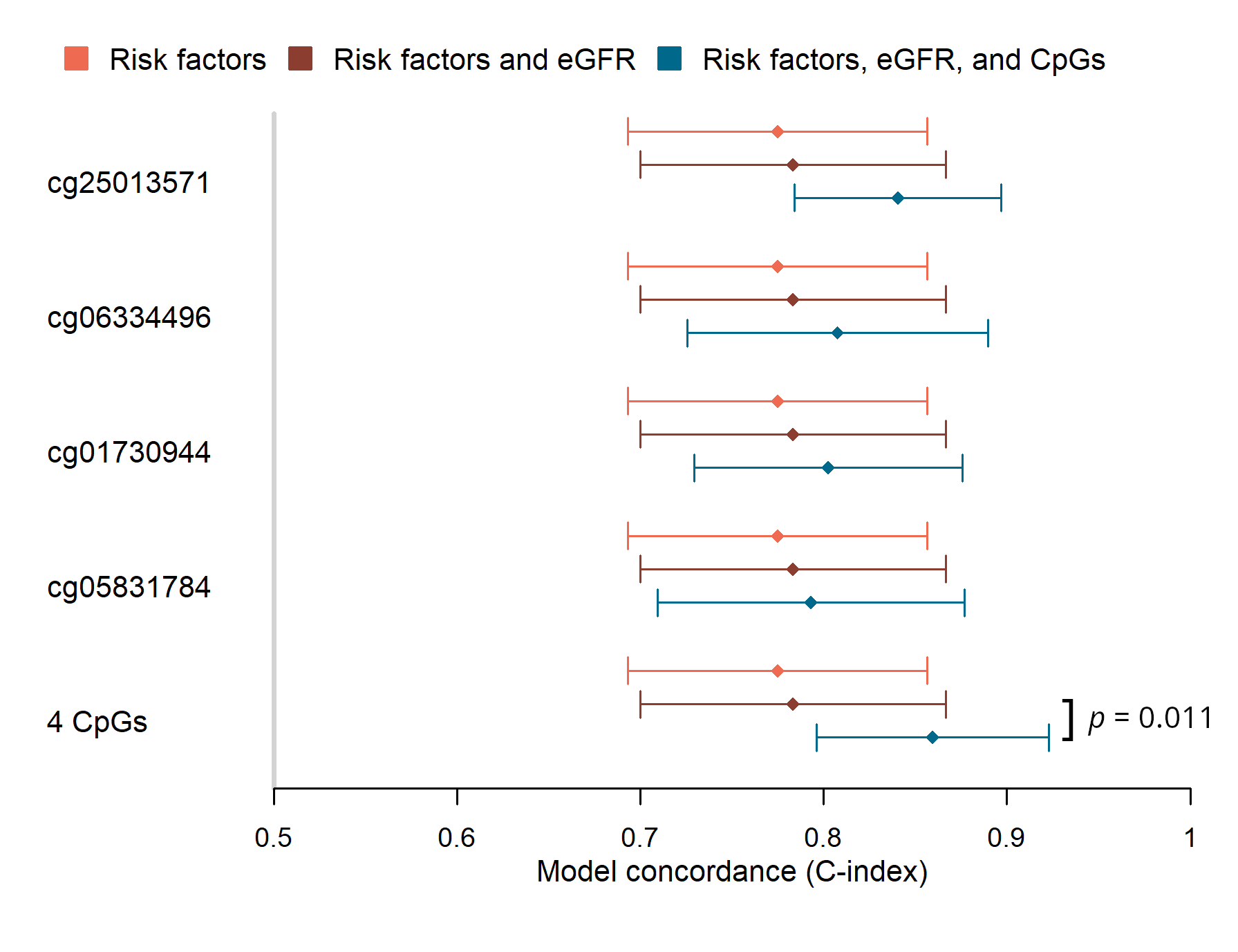

**Supplemental Figure 10.** Predictive power of early DKD progression associated CpGs. Concordances (C-index) and their 95% confidence intervals of different Cox proportional hazards model were plotted: “Risk factors” model (orange color) baseline triglyceride concentration, central obesity, and current smoking status in addition to age, sex, technical PCs 1–3 and mean methylation, as covariates. The “Risk factors and eGFR” model included additionally baseline eGFR, and it did not increase the predictive power of the model. The third model included the CpG sites separately or all four sites. When comparing the concordances (predictive power of the models), the only significant difference (*P*<0.05) is marked in the figure.

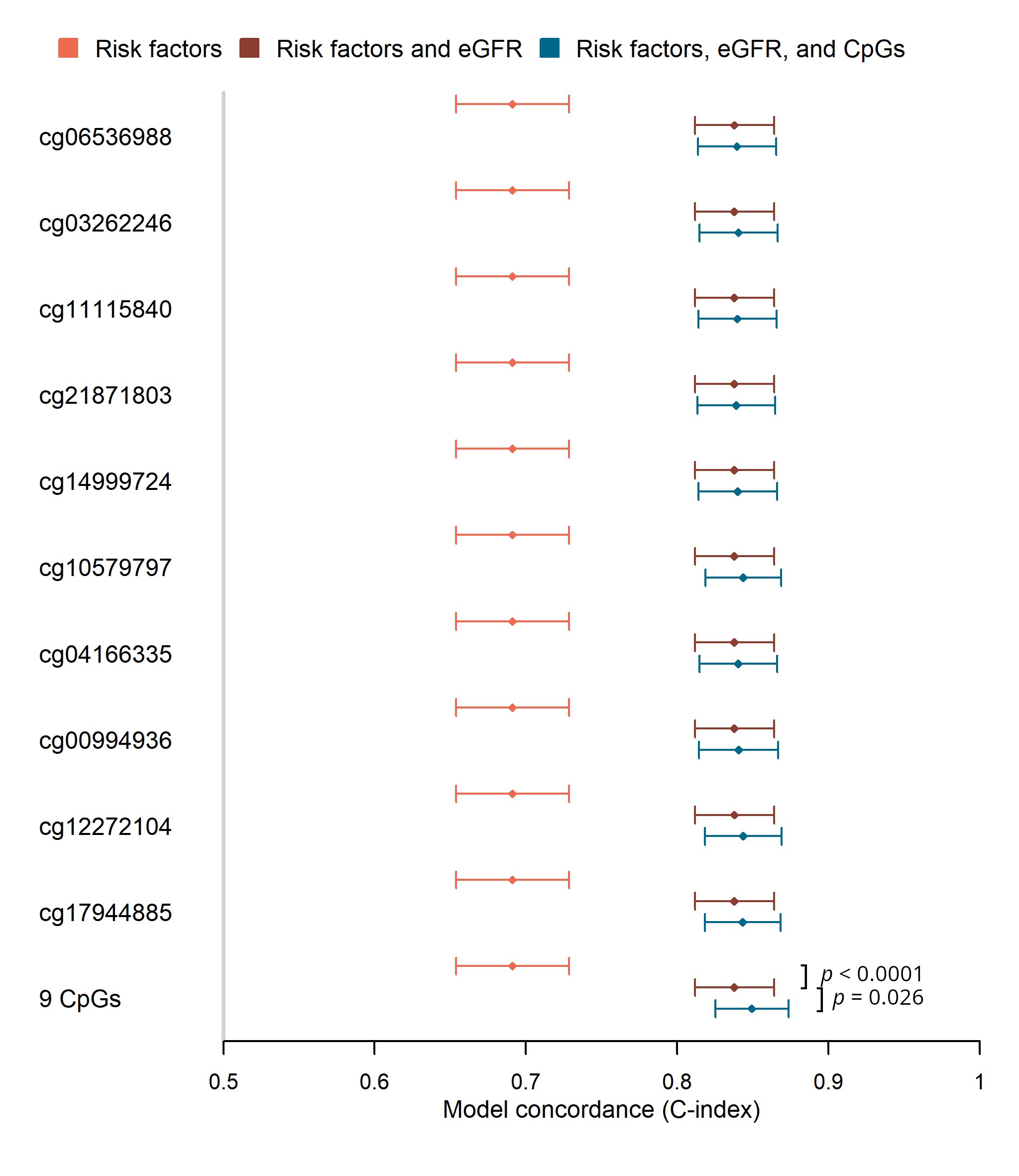

**Supplemental Figure 11.** Predictive power of late DKD progression associated CpGs. The diamonds show the concordance (C-index) and its 95% confidence intervals of three Cox proportional-hazards models applied for the early (*n*=363 with non-missing values in variables. *P*-values denote the significance of the increase in concordance compared to the previous model. Only significant differences (*P*<0.05) are marked in the figure. The “Risk factors” model (orange color) included triglyceride concentration, HbA_1c_, systolic blood pressure, along with six white blood cell proportions, technical PCs 1–3, mean methylation value, age, and sex. The second model (red color) included additionally baseline eGFR. The third model included methylation M values of nine (late DKD progression) significant CpGs, either separately or when combined.

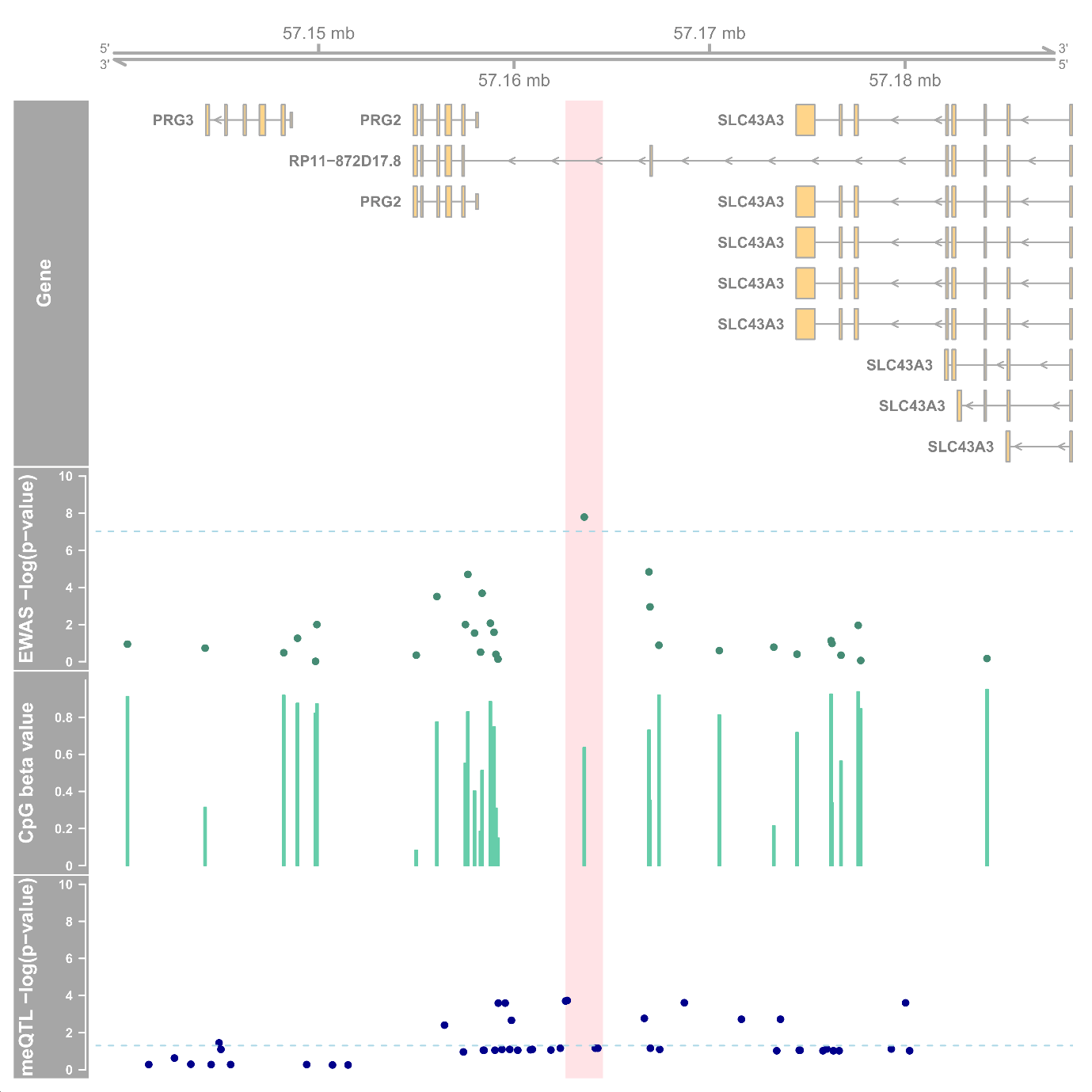

**Supplemental Figure 12**. Chromosome 11 region around cg14999724. The light red vertical highlight shows a 2,000 bp region around the top CpG cg14999724. Genes-track shows the UCSC genes and the EWAS -log10-transfromed *P*-value is from the late DKD progression EWAS of 373 individuals. Dashed blue line shows the epigenome-wide significance (*P*<9.4×10^–8^). The CpG beta values are mean methylation values in the late DKD progression cohort. The lowest track shows FDR-corrected *p*-values from the meQTL analysis in the FinnDiane cohort (*n*=765), where we tried to identify genetic variants associated with cg14999724 methylation. The top meQTL rs555097 was located –872 base pairs from the methylation site. Figure generated using the *Gviz* R-package.

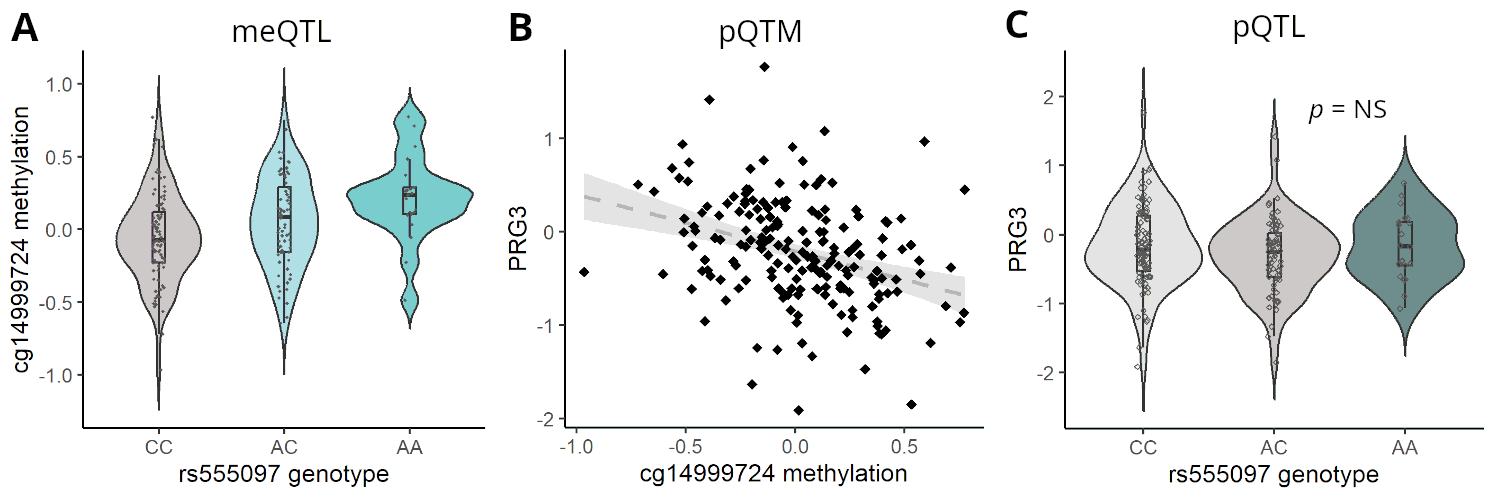

**Supplemental Figure 13**. CpG cg14999724 locus protein and SNV associations in 188 individuals with normal AER **A)** Violin plot shows SNV rs555097 association with methylation levels of cg14999724 (residualised methylation M-values values where blood cell variability and technical variability regressed is out. Of note, this SNV was a significant *cis-*meQTL for cg14999724 in the FinnDiane cohort analysis (*n*=765) and was significant in this normal AER sub-cohort analysis **B**) Methylation site cg14999724 was a significant *cis*-pQTM for serum PRG3 protein levels (beta=–0.18, *P*=1.7×10^–5^). Residualised methylation M values plotted (x-axis) with protein NPX values. **C**) The rs555097 genotypes were not significantly associated with PRG3 levels.

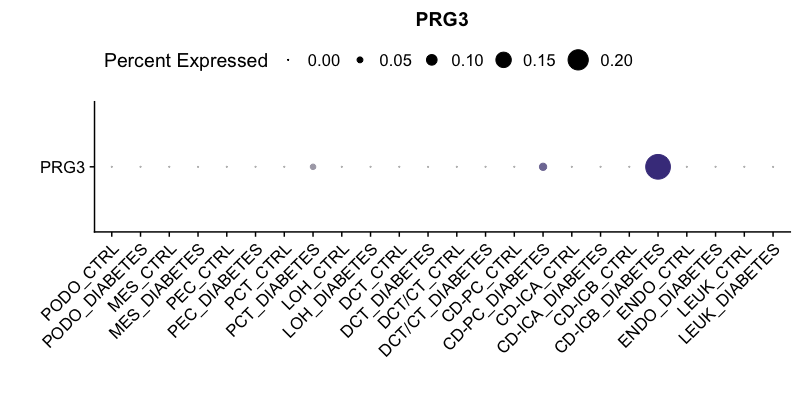

**Supplemental Figure 14.** PRG3 expression in human kidney single cell data set. Figure shows the 12 kidney single types in individuals with diabetes or without diabetes (CTRL) (Willson et al data accessed through Humphreys’ Lab browser at <http://humphreyslab.com>). Abbreviations PODO=podocytes; MESE=mesenchyme; PEC=parietal epithelial cell; PCT=proximal convoluted tubule; LOH=loop od Henle; DCT/CT=distal convoluted tubule/connecting tubule; CD-PC=collecting duct – principal cell; CD-IDA=collecting duct – intercalated cells A; CD-ICB=collecting duct – intercalated cells B; ENDO=endothelia; Leuk=leukocytes.

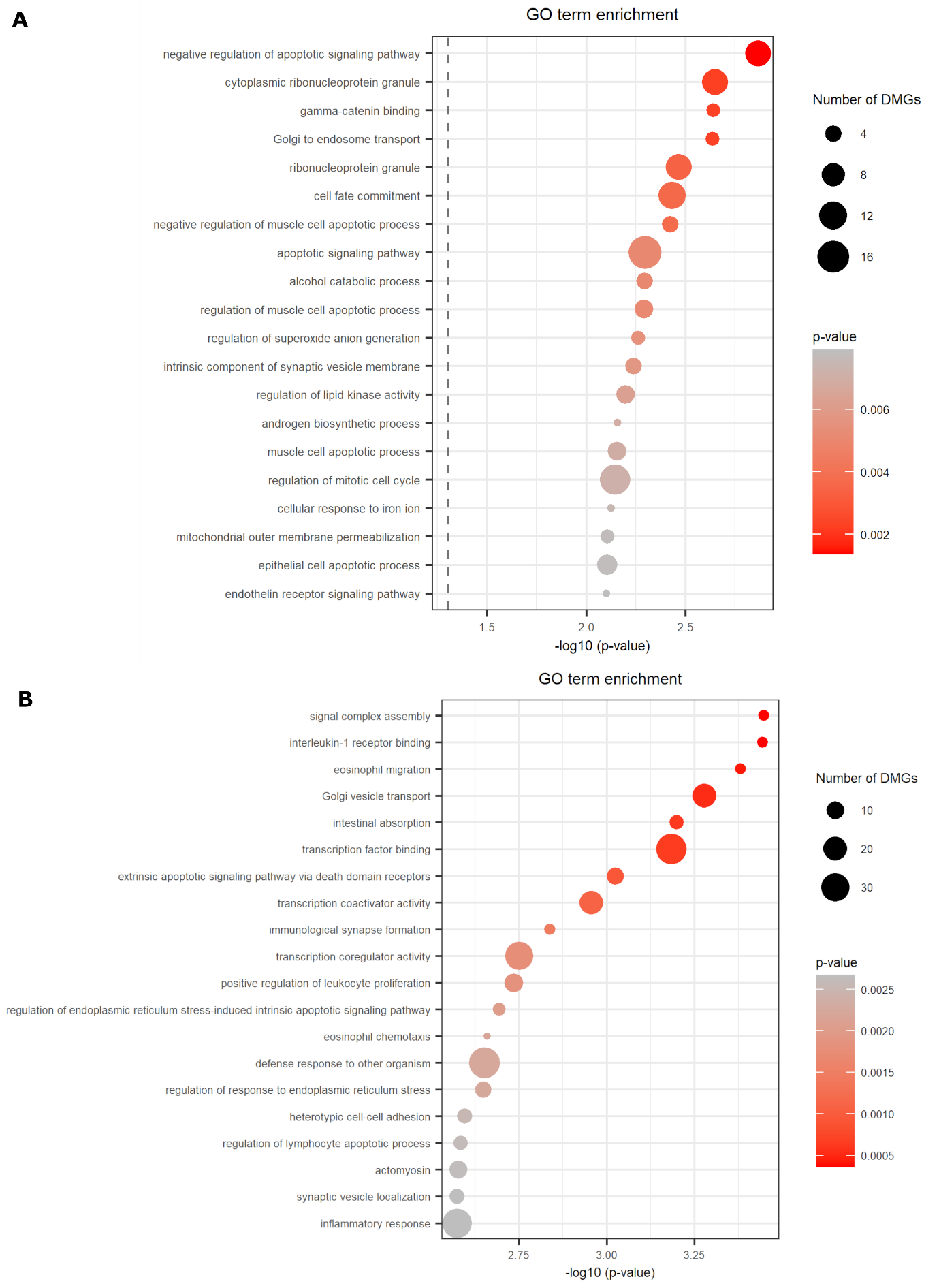

**Supplemental Figure 15**. Gene Ontology (GO) term enrichment results of the genes related to the early and late DKD progression –associated CpGs (*P*<10^–4^). A) GO term enrichment of genes related to the 317 early DKD progression –associated CpGs (due to a high overlap of top signals, results from the eGFR-adjusted and non-adjusted EWASs were combined) B) Genes related to the 750 late DKD progression –associated CpGs (model not adjusted for the baseline eGFR). Of note, no enrichment result was significant at FDR=0.05.

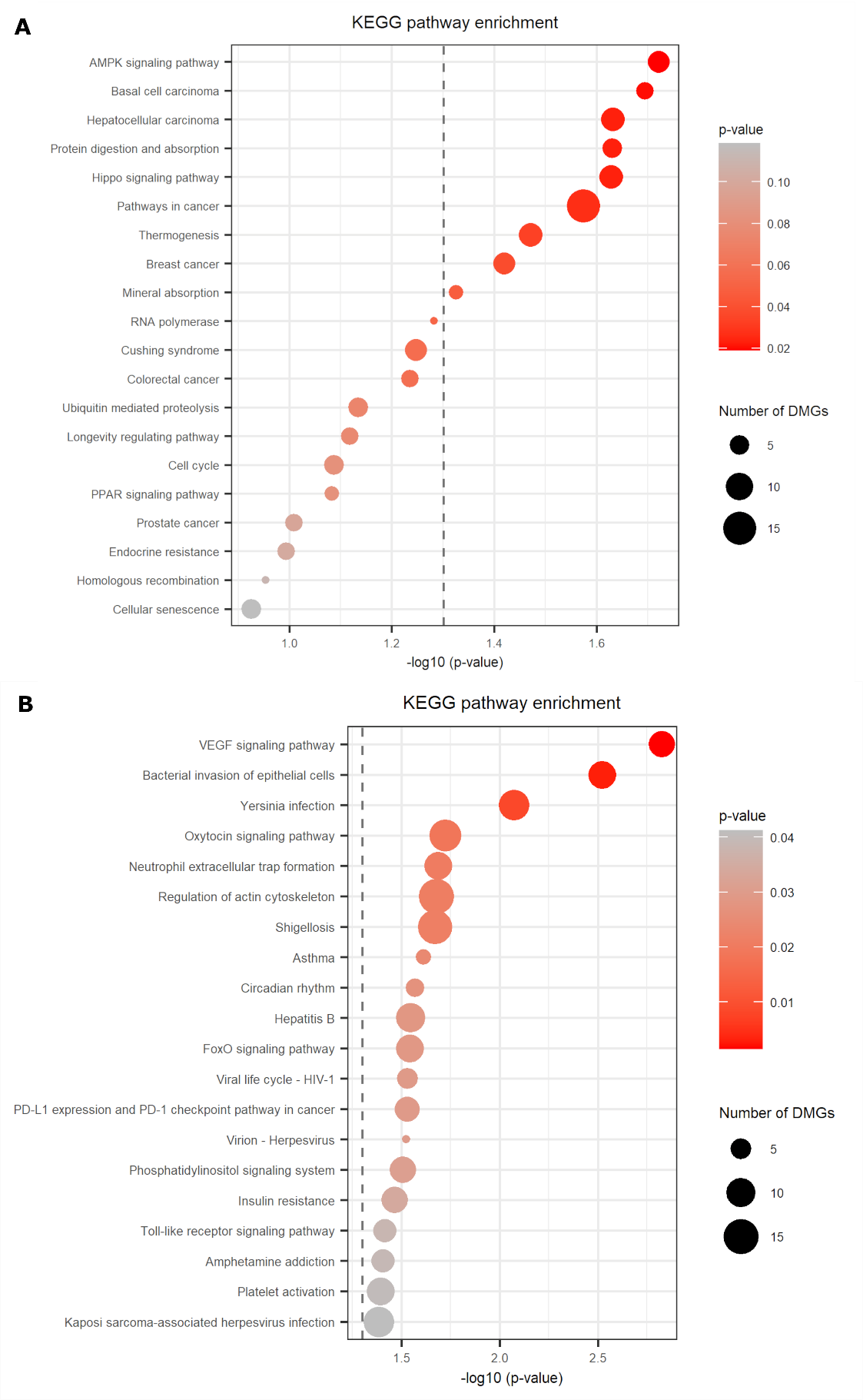

**Supplemental Figure 16**. KEGG pathway enrichment results of the genes related to the early and late DKD progression –associated CpGs (*P*<10^–4^). A) Enriched KEGG pathways for the genes related to the 317 early DKD progression –associated CpGs and B) 750 late DKD progression –associated CpGs. Dashed vertical line shows the P=0.05 level. Of note, no enrichment result was significant at FDR=0.05.

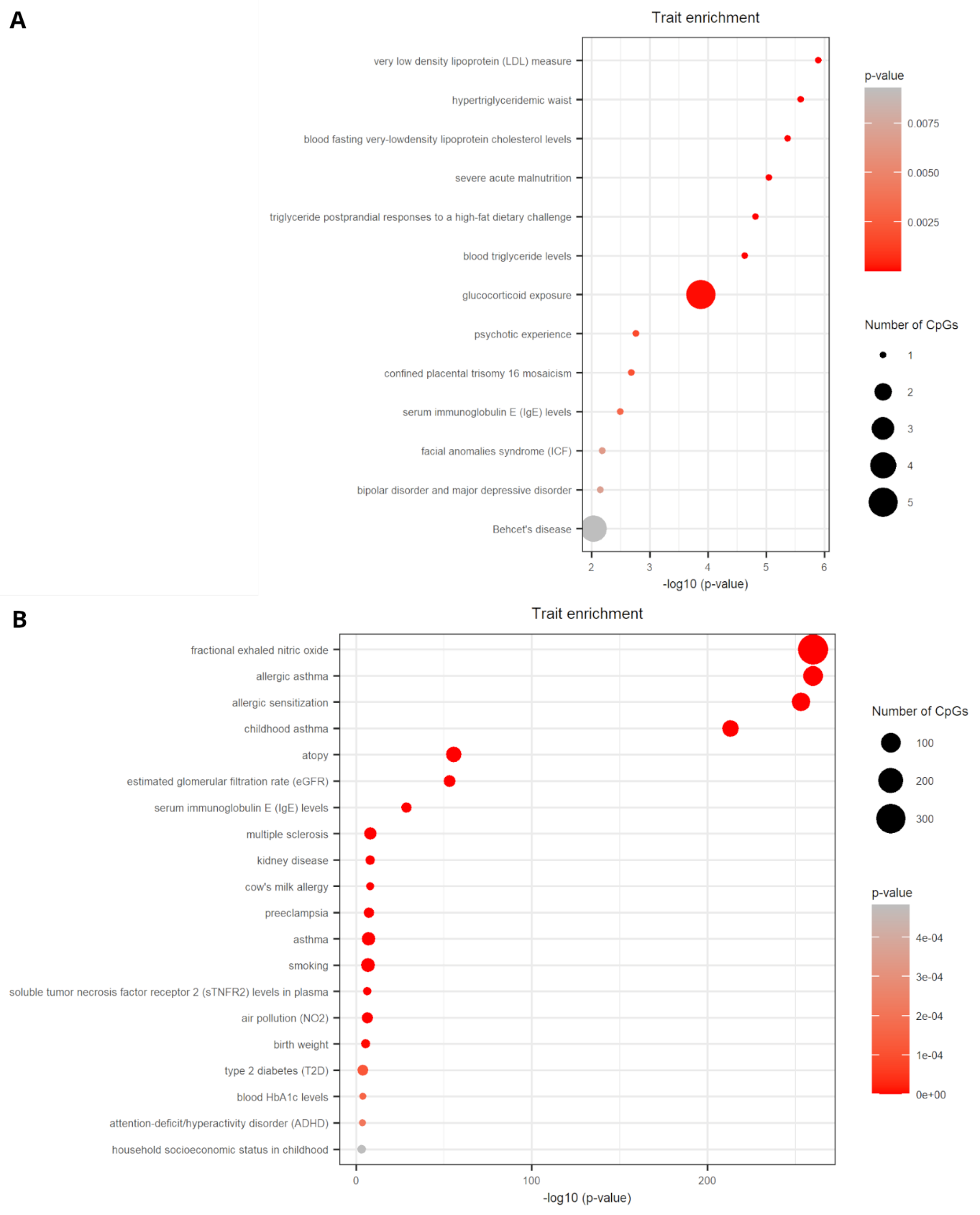

**Supplemental Figure 17.** Enrichment of CpGs associated with early and late DKD progression in traits with EWAS results in EWAS Atlas. Altogether A) 317 CpGs (early DKD progression associated) and B) 750 CpGs (late DKD progression associated) were analyzed. Trait enrichment analysis was carried out using EWAS Toolkit at <https://ngdc.cncb.ac.cn/ewas/toolkit>. Traits with ≥5 overlapping CpGs with our data were considered robust, i.e., excluding the other, mainly lipid related traits for early DKD progression (see panel A).

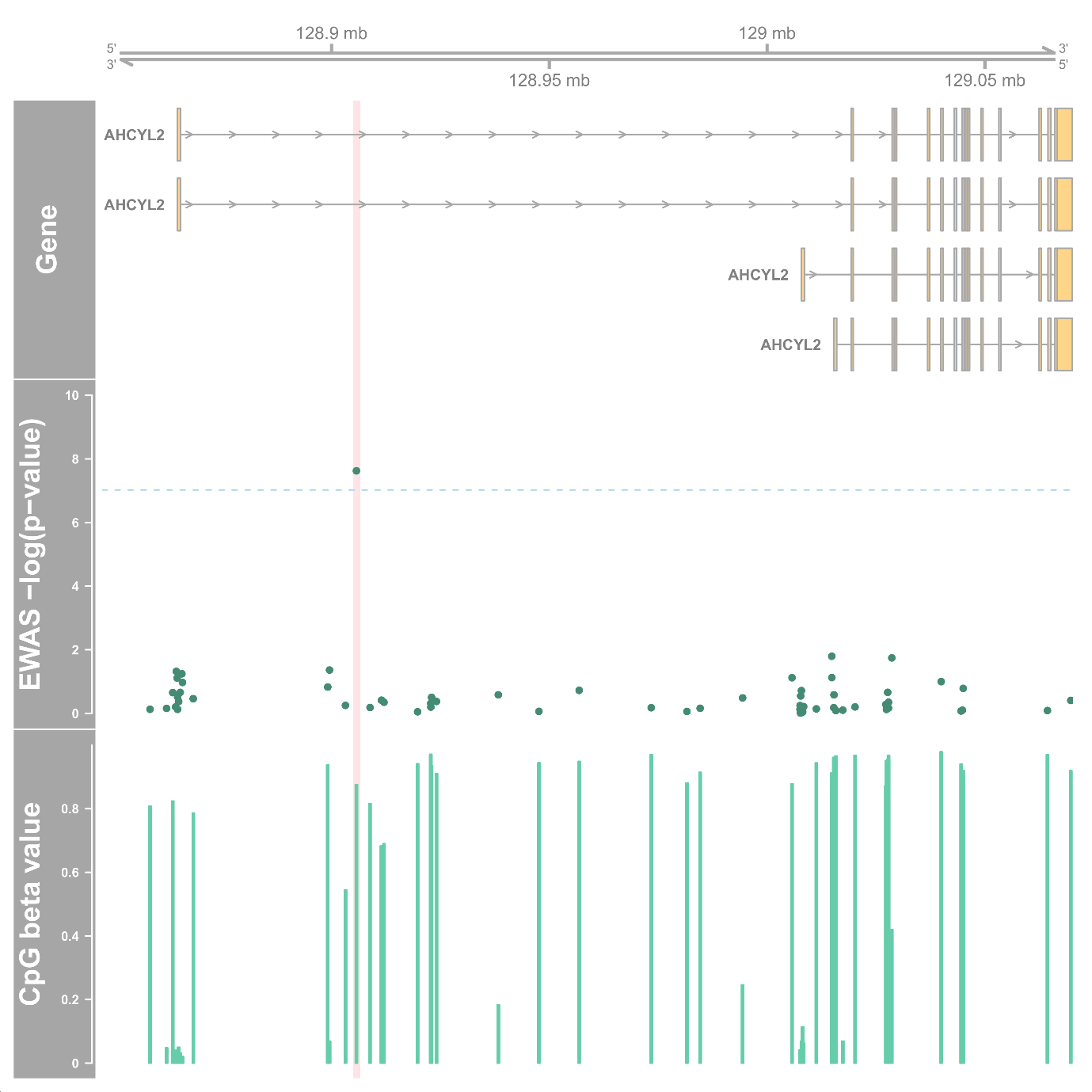

**Supplemental Figure 18**. Chromosome 7 region around CpG cg21871803. The light red vertical highlight shows a 2,000 bp region around the top CpG cg21871803. Genes-track shows the UCSC genes and the *P*-value is from the late DKD progression EWAS of 373 individuals. Dashed blue line shows the epigenome-wide significance (*P*<9.42×10^–8^). The CpG beta values are mean methylation values in the late DKD progression cohort.

#### **Supplemental Figure 19.** Physicians and nurses at the Finnish Diabetic Nephropathy (FinnDiane) study sites

| **FinnDiane Study Center** | **Physicians and nurses** |
| --- | --- |
| Anjalankoski Health Center | S.Koivula, T.Uggeldahl |
| Central Finland Central Hospital, Jyväskylä | T.Forslund, A.Halonen, A.Koistinen, P.Koskiaho, M.Laukkanen, J.Saltevo, M.Tiihonen |
| Central Hospital of Åland Islands, Mariehamn | M.Forsen, H.Granlund, A.-C.Jonsson, B.Nyroos |
| Central Hospital of Kanta-Häme, Hämeenlinna | P.Kinnunen, A.Orvola, T.Salonen, A.Vähänen |
| Central Hospital of Kymenlaakso, Kotka | R.Paldanius, M.Riihelä, L.Ryysy |
| Central Hospital of Länsi-Pohja, Kemi | H.Laukkanen, P.Nyländen, A.Sademies |
| Central Ostrobothnian Hospital District, Kokkola | S.Anderson, B.Asplund, U.Byskata, P.Liedes, M.Kuusela, T.Virkkala |
| City of Espoo Health Center: |  |
| Espoonlahti | A.Nikkola, E.Ritola |
| Tapiola | M.Niska, H.Saarinen |
| Samaria | E.Oukko-Ruponen, T.Virtanen |
| Viherlaakso | A.Lyytinen |
| City of Helsinki Health Center: |  |
| Puistola | H.Kari, T.Simonen |
| Suutarila | A.Kaprio, J.Kärkkäinen, B.Rantaeskola |
| Töölö | P.Kääriäinen, J.Haaga, A-L.Pietiläinen |
| City of Hyvinkää Health Center | S.Klemetti, T.Nyandoto, E.Rontu, S.Satuli-Autere |
| City of Vantaa Health Center: |  |
| Korso | R.Toivonen, H.Virtanen |
| Länsimäki | R.Ahonen, M.Ivaska-Suomela, A.Jauhiainen |
| Martinlaakso | M.Laine, T.Pellonpää, R.Puranen |
| Myyrmäki | A.Airas, J.Laakso, K.Rautavaara |
| Rekola | M.Erola, E.Jatkola |
| Tikkurila | R.Lönnblad, A.Malm, J.Mäkelä, E.Rautamo |
| Heinola Health Center | P.Hentunen, J.Lagerstam |
| Helsinki University Hospital, Department of Medicine, Division of Nephrology | T.Claesson, A.Dufva, N.Elonen, M.Eriksson, J.Fagerudd, M.Feodoroff, D.Gordin, P.-H.Groop, O.Heikkilä, K.Hietala, S.Hägg-Holmberg, F.Jansson Sigfrids, M.Korolainen, J.Kytö, S.Lindh, H.Paajanen, K.Pettersson-Fernholm, K.Rimpeläinen, M.Rosengård-Bärlund, M.Rönnback, L.Salovaara, A.Sandelin, M.Saraheimo, S.Satuli-Autere, R.Simonsen, P.Smidtslund, L.Thorn, H.Tikkanen, J.Tuomikangas, A.Tynjälä, K.Uljala, T.Vesisenaho, J.Wadén, A.Ylinen |
| Herttoniemi Hospital, Helsinki | V.Sipilä |
| Hospital of Lounais-Häme, Forssa | T.Kalliomäki, J.Koskelainen, R.Nikkanen, N.Savolainen, H.Sulonen, E.Valtonen |
| Hyvinkää Hospital | L. Norvio, A.Hämäläinen |
| Iisalmi Hospital | E.Toivanen |
| Jokilaakso Hospital, Jämsä | A.Parta, I.Pirttiniemi |
| Jorvi Hospital, Helsinki University Central Hospital | S.Aranko, S.Ervasti, R.Kauppinen-Mäkelin, A.Kuusisto, T.Leppälä, K.Nikkilä, L.Pekkonen |
| Jyväskylä Health Center, Kyllö | K.Nuorva, M.Tiihonen |
| Kainuu Central Hospital, Kajaani | S.Jokelainen, K.Kananen, M.Karjalainen, P.Kemppainen, A-M.Mankinen, A.Reponen, M.Sankari |
| Kerava Health Center | H.Stuckey, P.Suominen |
| Kirkkonummi Health Center | A.Lappalainen, M.Liimatainen, J.Santaholma |
| Kivelä Hospital, Helsinki | A.Aimolahti, E.Huovinen |
| Koskela Hospital, Helsinki | V.Ilkka, M.Lehtimäki |
| Kotka Health Center | E.Pälikkö-Kontinen, A.Vanhanen |
| Kouvola Health Center | E.Koskinen, T.Siitonen |
| Kuopio University Hospital | E.Huttunen, R.Ikäheimo, P.Karhapää, P.Kekäläinen, M.Laakso, T.Lakka, E.Lampainen, L.Moilanen, S. Tanskanen, L.Niskanen, U.Tuovinen, I.Vauhkonen, E.Voutilainen |
| Kuusamo Health Center | T.Kääriäinen, E.Isopoussu |
| Kuusankoski Hospital | E.Kilkki, I.Koskinen, L.Riihelä |
| Laakso Hospital, Helsinki | T.Meriläinen, P.Poukka, R.Savolainen, N.Uhlenius |
| Lahti City Hospital | A.Mäkelä, M.Tanner |
| Lapland Central Hospital, Rovaniemi | L.Hyvärinen, K.Lampela, S.Pöykkö, T.Rompasaari, S.Severinkangas, T.Tulokas |
| Lappeenranta Health Center | P. Erola, L.Härkönen, P.Linkola, T.Pekkanen, I.Pulli, E.Repo |
| Lohja Hospital | T.Granlund, K.Hietanen, M.Porrassalmi, M.Saari, T.Salonen, M.Tiikkainen, |
| Länsi-Uusimaa Hospital, Tammisaari | I.-M.Jousmaa, J.Rinne |
| Loimaa Health Center | A.Mäkelä, P.Eloranta |
| Malmi Hospital, Helsinki | H.Lanki, S.Moilanen, M.Tilly-Kiesi |
| Mikkeli Central Hospital | A.Gynther, R.Manninen, P.Nironen, M.Salminen, T.Vänttinen |
| Mänttä Regional Hospital | I.Pirttiniemi, A-M.Hänninen |
| North Karelian Hospital, Joensuu | U-M.Henttula, P.Kekäläinen, M.Pietarinen, A.Rissanen, M.Voutilainen |
| Nurmijärvi Health Center | A.Burgos, K.Urtamo |
| Oulaskangas Hospital, Oulainen | E.Jokelainen, P-L.Jylkkä, E.Kaarlela, J.Vuolaspuro |
| Oulu Health Center | L.Hiltunen, R.Häkkinen, S.Keinänen-Kiukaanniemi |
| Oulu University Hospital | R.Ikäheimo |
| Päijät-Häme Central Hospital | H.Haapamäki, A.Helanterä, S.Hämäläinen, V.Ilvesmäki, H.Miettinen |
| Palokka Health Center | P.Sopanen, L.Welling |
| Pieksämäki Hospital | V.Sevtsenko, M.Tamminen |
| Pietarsaari Hospital | M-L.Holmbäck, B.Isomaa, L.Sarelin |
| Pori City Hospital | P.Ahonen, P.Merisalo, E.Muurinen, K.Sävelä |
| Porvoo Hospital | M.Kallio, B.Rask, S.Rämö |
| Raahe Hospital | A.Holma, M.Honkala, A.Tuomivaara, R.Vainionpää |
| Rauma Hospital | K.Laine, K.Saarinen, T.Salminen |
| Riihimäki Hospital | P.Aalto, E.Immonen, L.Juurinen |
| Salo Hospital | A.Alanko, J.Lapinleimu, P.Rautio, M.Virtanen |
| Satakunta Central Hospital, Pori | M.Asola, M.Juhola, P.Kunelius, M.-L.Lahdenmäki, P.Pääkkönen, M.Rautavirta |
| Savonlinna Central Hospital | T.Pulli, P.Sallinen, M.Taskinen, E.Tolvanen, T.Tuominen, H.Valtonen, A.Vartia, S-L.Viitanen |
| Seinäjoki Central Hospital | O.Antila, E.Korpi-Hyövälti, T.Latvala, E.Leijala, T.Leikkari, M.Punkari N.Rantamäki, H.Vähävuori |
| South Karelia Central Hospital, Lappeenranta | T.Ensala, E.Hussi, R.Härkönen, U.Nyholm, J.Toivanen |
| Tampere Health Center | A.Vaden, P.Alarotu, E.Kujansuu, H.Kirkkopelto-Jokinen, M.Helin, S.Gummerus, L.Calonius, T.Niskanen, T.Kaitala, T.Vatanen |
| Tampere University Hospital | P. Hannula, I.Ala-Houhala, R.Kannisto, T.Kuningas, P.Lampinen, M.Määttä,H.Oksala, T.Oksanen, A.Putila, H.Saha, K.Salonen, H.Tauriainen, S.Tulokas |
| Tiirismaa Health Center, Hollola | T.Kivelä, L.Petlin, L.Savolainen |
| Turku Health Center | A.Artukka, I.Hämäläinen, L.Lehtinen, E.Pyysalo, H.Virtamo, M.Viinikkala, M.Vähätalo |
| Turku University Central Hospital | K.Breitholz, R.Eskola, K.Metsärinne, U.Pietilä, P.Saarinen, R.Tuominen, S.Äyräpää |
| Vaajakoski Health Center | K.Mäkinen, P.Sopanen |
| Valkeakoski Regional Hospital | S.Ojanen, E.Valtonen, H.Ylönen, M.Rautiainen, T.Immonen |
| Vammala Regional Hospital | I.Isomäki, R.Kroneld, L.Mustaniemi, M.Tapiolinna-Mäkelä |
| Vasa Central Hospital | S.Bergkulla, U.Hautamäki, V-A.Myllyniemi, I.Rusk |

#### **Supplemental Figure 20**. Members of the GENIE Consortium.

| **Name** | **Affiliations** |
| --- | --- |
| **Massachusetts General Hospital and Broad Institute, Boston, MA, USA** | |
| Joel N Hirschhorn | Programs in Metabolism and Medical & Population Genetics, Broad Institute, Cambridge, MA USA.  Division of Endocrinology, Boston Children’s Hospital, Boston, MA, USA  Department of Pediatrics and Genetics, Harvard Medical School, Boston, MA, USA |
| Jose C Florez | Programs in Metabolism and Medical & Population Genetics, Broad Institute, Cambridge, MA USA  Diabetes Unit and Center for Genomic Medicine, Massachusetts General Hospital, Boston, MA USA.  Department of Medicine, Harvard Medical School, Boston, MA USA. |
| Raymond Kreienkamp | Division of Endocrinology, Boston Children’s Hospital, Boston, MA, USA Diabetes Unit and Center for Genomic Medicine, Massachusetts General Hospital, Boston, MA USA. |
| **The FinnDiane Study Group, Folkhälsan Research Center, Helsinki, Finland** | |
| Emma H Dahlström | Folkhälsan Institute of Genetics, Folkhälsan Research Center, Helsinki, Finland.  Department of Nephrology, University of Helsinki and Helsinki University Hospital, Helsinki, Finland.  Research Program for Clinical and Molecular Metabolism, Faculty of Medicine, University of Helsinki, 00290, Helsinki, Finland. |
| Anna Syreeni | Folkhälsan Institute of Genetics, Folkhälsan Research Center, Helsinki, Finland.  Department of Nephrology, University of Helsinki and Helsinki University Hospital, Helsinki, Finland.  Research Program for Clinical and Molecular Metabolism, Faculty of Medicine, University of Helsinki, 00290, Helsinki, Finland. |
| Erkka Valo | Folkhälsan Institute of Genetics, Folkhälsan Research Center, Helsinki, Finland.  Department of Nephrology, University of Helsinki and Helsinki University Hospital, Helsinki, Finland.  Research Program for Clinical and Molecular Metabolism, Faculty of Medicine, University of Helsinki, 00290, Helsinki, Finland. |
| Valma Harjutsalo | Folkhälsan Institute of Genetics, Folkhälsan Research Center, Helsinki, Finland.  Department of Nephrology, University of Helsinki and Helsinki University Hospital, Helsinki, Finland.  Research Program for Clinical and Molecular Metabolism, Faculty of Medicine, University of Helsinki, 00290, Helsinki, Finland. |
| Per-Henrik Groop | Folkhälsan Institute of Genetics, Folkhälsan Research Center, Helsinki, Finland.  Department of Nephrology, University of Helsinki and Helsinki University Hospital, Helsinki, Finland.  Research Program for Clinical and Molecular Metabolism, Faculty of Medicine, University of Helsinki, 00290, Helsinki, Finland.  Department of Diabetes, Central Clinical School, Monash University, Melbourne, Victoria, Australia. |
| Niina Sandholm | Folkhälsan Institute of Genetics, Folkhälsan Research Center, Helsinki, Finland.  Department of Nephrology, University of Helsinki and Helsinki University Hospital, Helsinki, Finland.  Research Program for Clinical and Molecular Metabolism, Faculty of Medicine, University of Helsinki, 00290, Helsinki, Finland. |
| **Queen's University Belfast, Belfast, Northern Ireland** | |
| Laura J Smyth | Molecular Epidemiology Research Group, Centre for Public Health, Queen's University Belfast, Belfast, UK. |
| Katie Kerr | Molecular Epidemiology Research Group, Centre for Public Health, Queen's University Belfast, Belfast, UK. |
| Jill Kilner | Molecular Epidemiology Research Group, Centre for Public Health, Queen's University Belfast, Belfast, UK. |
| Yogesh Gupta | Molecular Epidemiology Research Group, Centre for Public Health, Queen's University Belfast, Belfast, UK. |
| Claire Hill | Molecular Epidemiology Research Group, Centre for Public Health, Queen's University Belfast, Belfast, UK. |
| Christopher Wooster | Molecular Epidemiology Research Group, Centre for Public Health, Queen's University Belfast, Belfast, UK. |
| Kerry Anderson | Molecular Epidemiology Research Group, Centre for Public Health, Queen's University Belfast, Belfast, UK. |
| Gareth J McKay | Molecular Epidemiology Research Group, Centre for Public Health, Queen's University Belfast, Belfast, UK. |
| Amy Jayne McKnight | Molecular Epidemiology Research Group, Centre for Public Health, Queen's University Belfast, Belfast, UK. |
| Alexander P Maxwell | Molecular Epidemiology Research Group, Centre for Public Health, Queen's University Belfast, Belfast, UK.  Regional Nephrology Unit, Belfast City Hospital, Belfast, Northern Ireland, UK. |
| **Diabetes Complications Research Centre, University College Dublin, Dublin Ireland** | |
| Ciarán Kennedy | Diabetes Complications Research Centre, Conway Institute, School of Medicine, University College Dublin, Dublin Ireland. |
| Ross Doyle | Diabetes Complications Research Centre, Conway Institute, School of Medicine, University College Dublin, Dublin Ireland. |
| Eoin Brennan | Diabetes Complications Research Centre, Conway Institute, School of Medicine, University College Dublin, Dublin Ireland. |
| Darrell Andrews | Diabetes Complications Research Centre, Conway Institute, School of Medicine, University College Dublin, Dublin Ireland. |
| Denise Sadlier | Mater Misericordiae Hospital, Dublin, Ireland D07 K201. |
| Finian Martin | Diabetes Complications Research Centre, Conway Institute, School of Medicine, University College Dublin, Dublin Ireland. |
| Catherine Godson | Diabetes Complications Research Centre, Conway Institute, School of Medicine, University College Dublin, Dublin Ireland. |
| **University of Michigan School of Medicine, Ann Arbor, MI, USA** | |
| Viji Nair | Department of Medicine-Nephrology, University of Michigan School of Medicine, Ann Arbor, MI 48109, USA. |
| Damian Fermin | Department of Pediatrics-Nephrology, University of Michigan School of Medicine, Ann Arbor, MI 48109, USA. |
| Lalita Subramanian | Department of Medicine-Nephrology, University of Michigan School of Medicine, Ann Arbor, MI 48109, USA. |
| Matthias Kretzler | Department of Internal Medicine, University of Michigan, Ann Arbor, Michigan, USA. |
| **University of Pennsylvania, Perelman School of Medicine, Philadelphia, PA, USA.** | |
| Hongbo Liu | Renal, Electrolyte, and Hypertension Division, Department of Medicine, University of Pennsylvania, Perelman School of Medicine, Philadelphia, PA, USA.  Institute for Diabetes, Obesity, and Metabolism, University of Pennsylvania, Perelman School of Medicine, Philadelphia, PA, USA.  Department of Genetics, University of Pennsylvania, Perelman School of Medicine, Philadelphia, PA, USA |
| Katalin Susztak | Renal, Electrolyte, and Hypertension Division, Department of Medicine, University of Pennsylvania, Perelman School of Medicine, Philadelphia, PA, USA.  Institute for Diabetes, Obesity, and Metabolism, University of Pennsylvania, Perelman School of Medicine, Philadelphia, PA, USA.  Department of Genetics, University of Pennsylvania, Perelman School of Medicine, Philadelphia, PA, USA |
| **University of California San Diego, La Jolla, CA, USA** | |
| Rany M Salem | Herbert Wertheim School of Public Health and Human Longevity Science, University of California San Diego, La Jolla, CA, USA |
| **University of Colorado School of Medicine, Aurora, CO, USA** | |
| Joanne B Cole | Department of Biomedical Informatics, University of Colorado School of Medicine, Aurora, CO, USA  Programs in Metabolism and Medical & Population Genetics, Broad Institute, Cambridge, MA USA.  Diabetes Unit and Center for Genomic Medicine, Massachusetts General Hospital, Boston, MA USA. |
